# The circulating blood proteome of childhood acute leukemia

**DOI:** 10.64898/2026.06.17.26355844

**Authors:** Anna Pia Enblad, Maria A. Globisch, Dea Gogishvili, Tiina Tuononen, Olga Krali, Anders Lundmark, Laura Oksa, Charlotte Hjort, Mariya Lysenkova Wiklander, Linda Holmfeldt, Mikael Åberg, Josefine Palle, Signe Modvig, Olli Lohi, Merja Heinäniemi, Arja Harila, Jessica Nordlund

## Abstract

The circulating blood proteome provides a systemic readout of disease biology and holds promise for advancing diagnostics and disease monitoring in pediatric leukemia. Here, we profiled 3072 proteins in diagnostic serum from 54 children with acute lymphoblastic leukemia (ALL), 21 with acute myeloid leukemia (AML), and 12 healthy controls using the Olink Proximity Extension Assay. We observed profound alterations in circulating protein levels in leukemia patients compared with controls and identified immunophenotype-specific proteins, including SIGLEC15 in B-cell precursor ALL (BCP-ALL), NOTCH1 in T-ALL, and CEBPA in AML, all which remained high even in patients with low (<20%) or no peripheral blood blasts. Within BCP-ALL, molecular subtypes were reflected in the circulating proteome; for example, DSC2 and PTPRK were elevated in *ETV6*::*RUNX1*-positive cases, while IL-6R and ADAM8 were higher in High Hyperdiploid cases. Angiogenic growth factors decreased across all leukemia patients compared with controls, suggesting a fragile peripheral vasculature at diagnosis. Integration with external datasets revealed the likely cellular source of abundant proteins and examination of an external cohort validated our subtype-specific findings. Together, these results define shared and distinct proteomic signatures across pediatric acute leukemias and highlight candidate biomarkers for diagnostics and disease monitoring.

**Graphical abstract:** 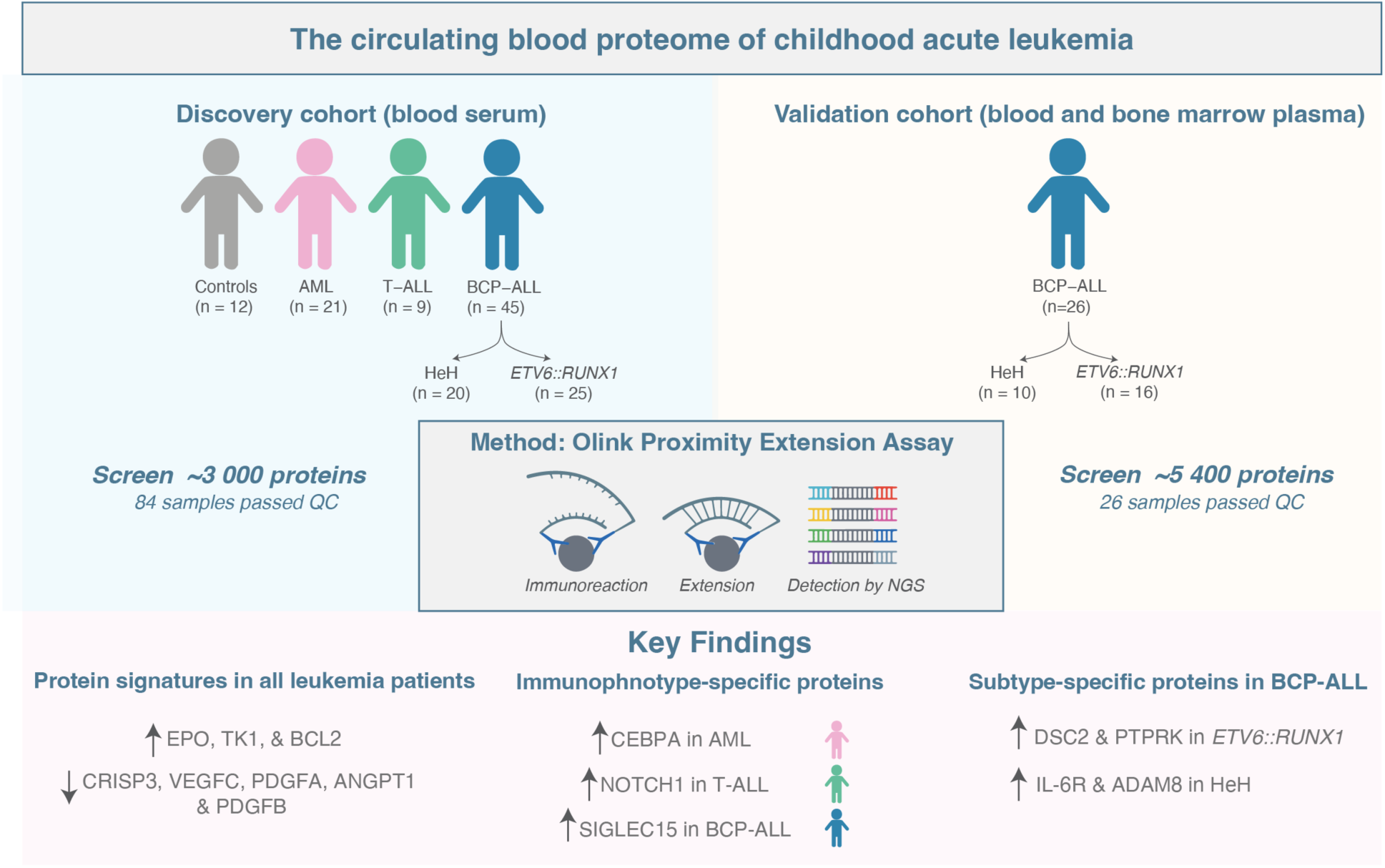

## Introduction

Acute leukemia is the most common childhood malignancy worldwide. Advances in risk-adapted treatments have raised the survival rates for children with acute lymphoblastic leukemia (ALL) to above 90%(1, 2), yet outcomes remain poorer for acute myeloid leukemia (AML), where long-term remission is achieved in only ∼70% of cases (3). Genomics-based diagnostics have transformed leukemia classification and informed tailored treatment approaches (4–8), but these methods only provide a blast-intrinsic perspective. Patients’ outcomes are also shaped by systemic physiological responses (9), which remain less well understood. The importance of the immune system in modulating treatment responses has become increasingly evident, particularly for immunotherapies such as chimeric antigen receptor T-cell therapy and bi-specific T-cell engagers like Blinatumomab (10, 11). To fully harness these approaches, biomarkers that capture both blast-characteristics and host response are needed.

The circulating blood proteome offers a minimally invasive readout of disease status and encompasses both immune cell activity as well as blast-immune system interactions (12). High-throughput platforms such as the proximity extension assay (PEA), now enable profiling of thousands of proteins from small blood volumes (13–16). While leukemic cell proteomes are well characterized (17, 18), the circulating blood proteome in pediatric leukemia remains underexplored. Previous ALL studies have focused on candidate proteins, within limited cohorts(19–25) and adult AML has been profiled extensively (26), however no systematic analysis has yet addressed circulating blood proteomes across immunophenotypic and molecular subtypes of pediatric leukemia.

This study applied a next-generation proteomics approach (PEA) to profile 3072 circulating proteins in diagnostic serum from children with ALL and AML. By integrating proteomic, immunophenotypic, and genetic subtype data, we aimed to identify signatures that reflect both blast-intrinsic features and systemic physiological responses.

## Materials and Methods

### Patient selection (discovery cohort)

Diagnostic serum samples from pediatric ALL and AML patients were obtained from the Nordic Society of Pediatric Hematology and Oncology (NOPHO) Leukemia Biobank in Uppsala, Sweden. In total, 75 acute leukemia samples were included. Clinical information (subtype, immunophenotype, as well as hemoglobin, leukocyte, blast, and platelet count at diagnosis) was retrieved from the Swedish Pediatric Cancer Registry (Stockholm, Sweden). For the B-cell precursor (BCP) ALL patients, subtype inclusion was limited to the two most common subtypes: High Hyperdiploidy (HeH) and *ETV6*::*RUNX1*. Samples were collected between the years 2006 and 2017.

Serum from 12 healthy children (siblings of pediatric oncology patients unrelated to this cohort) served as controls. These samples were collected between February 2022 and June 2023. Both acute leukemia samples and control samples were collected according to the same procedure. Briefly, blood was drawn in serum-separating tubes, allowed to clot, centrifuged, and stored at −80°C. None of the samples had been previously thawed. Informed consent was obtained from the children and/or their guardians for sample storage and future research use. The study was conducted in accordance with the Declaration of Helsinki and approved by the Swedish Ethical Review Authority [#2021-05213].

### Serum protein profiling and quality control (discovery cohort)

The serum samples (n=87) were thawed at room temperature, and 50μL of each was aliquoted into a 96-well plate in a randomized order to minimize potential batch effects. The plate was stored at −80°C until processing. Proteomic profiling was performed using the Olink Explore 3072 platform at SciLifeLab (Uppsala, Sweden) according to the manufacturer’s protocol. Details regarding the platform, assayed proteins, and sequencing parameters can be found in the **Supplemental Methods**.

Three samples failed quality control (QC) and were excluded (**Supplemental Table S1**). Proteins that failed to pass Olink’s batch release QC criteria or were below the limit of detection (LOD) were excluded, leaving 2897 proteins for downstream analyses (**Supplemental Table S2**).

### Publicly available cellular transcriptomics and proteomics datasets

Gene and protein expression data from pediatric patient-derived ALL and AML cells were obtained from published datasets (17, 18, 27). Bulk RNA-sequencing (RNA-seq) data from diagnostic BCP-ALL (n=46 cases with HeH and n=32 with the *ETV6*::*RUNX1* fusion) and T-ALL (n=19) samples were retrieved from the Gene Expression Omnibus (GEO; accession number GSE227832)(27). Of these, 3 HeH, 6 *ETV6::RUNX1*, and 2 T-ALL cases overlapped with the discovery cohort, while the remaining samples derived from independent patients. RNA-seq data from unmatched diagnostic AML samples (n=18) were obtained from the SciLifeLab Data Repository (https://doi.org/10.17044/scilifelab.13105229.v1) (18). The AML transcriptome dataset included the following set of subtypes; CEBPA-mutated (n=1), inv(16) (n=2), t(8;21) (n=3), trisomy 8 (n=1), WT1-mutated (n=1), FLT3-ITD (n=5) and unknown (n=5). Details regarding RNA-seq data processing are provided in the **Supplemental Methods**. Differential protein expression data from unmatched diagnostic pediatric BCP-ALL samples with HeH (n= 48) or *ETV6*::*RUNX1* (n= 41) subtypes were obtained from a publicly available mass spectrometry (MS) dataset (17). A summary of patient characteristics for the MS and transcriptomic cohorts is provided in **Supplemental Table S3**.

### Statistical analyses

All analyses were performed in RStudio (version 2024.04.2) or Python (version 3.11.9). Principal component analysis (PCA) plots were generated using the *Olink Analyze* R package. Differential protein expression was assessed using *limma* (28). The Benjamini-Hochberg method was used to adjust for multiple testing. Age, sex, and, when applicable, molecular subtype were included as covariates. Statistical significance was defined as a false discovery rate (FDR)-adjusted p-value < 0.05 and absolute Log2 fold change (|Log2FC|) > 1.5. A total of five comparisons were made: first all leukemia samples were pooled and compared with controls then the three immunophenotypes (AML, T-ALL and BCP-ALL) were independently compared with controls, and lastly within the BCP-ALL group HeH patients were compared with *ETV6*::*RUNX1-*positive samples.

Pathway enrichment was evaluated using the *clusterProfiler* package; we employed both gene set enrichment analysis (GSEA) and an over-representation analysis ORA to comprehensively capture both coordinated pathway-level shifts and pathways with strongly dysregulated individual proteins. The Molecular Signatures Database hallmark gene sets were used for annotation.

For the RNA-seq data, differential expression analysis was performed using *limma* (threshold = FDR < 0.05 and |Log2FC| > 1.5). Spearman pairwise correlations were calculated between the mean expression value levels or Log2FC value of proteins and the corresponding gene expression (GEX) values from RNA-seq.

For proteins with detectable mRNA expression in blast cells (Log2(CPM) > 1), Pearson correlations were calculated between Olink Log2FC and corresponding mRNA expression Log2FC. Similarly, for proteins detected in both serum and cells, correlations were computed between Olink and mass spectrometry-derived cellular protein abundance Log2FC.

### Clinical characteristics

To assess the relationship between overall protein abundance and patients’ peripheral blood counts, we calculated the median NPX value across all measured proteins for each patient sample. White blood cell (WBC) and platelet counts at diagnosis were log-transformed. Pearson correlations were calculated between median NPX and each clinical variable then stratified by immunophenotype, as well as between individual protein abundance values and clinical variables.

### Plasma protein profiling (validation cohort)

Validation was performed using peripheral blood plasma (HeH n=7, *ETV6*::*RUNX1* n=10) and bone marrow plasma (HeH n=10, *ETV6*::*RUNX1* n=16) collected at diagnosis from pediatric B-ALL patients with confirmed molecular subtypes. Protein profiling was performed in two batches using the Olink HT 5072 platform at SciLifeLab (Uppsala, Sweden) according to the manufacturer’s protocol. Experimental details regarding the plasma collection, sequencing parameters, data processing, batch correction and statical analyses can be found in the **Supplemental Methods**. Samples from the validation cohort were collected under the ethical permit #R13109 and approved by the Pirkanmaa Hospital District Ethical Committee in Finland.

### Protein annotations

The cellular localizations of differentially expressed proteins were retrieved from UniProt via the *mapUniProt* function in the *UniProt.ws* package, utilizing the UniProt API. Proteins annotated as “secreted” in UniProt were classified as extracellular, which took precedence over other locations (e.g., VEGFR2, a membrane receptor with a secreted form, was labeled extracellular). Proteins were categorized as membrane-associated if annotated only as “membrane,” and as intracellular if annotated as cytoplasmic, nuclear or other internal compartments. Proteins without clear annotations were labeled as unclassified. Protein localization annotations were extended using DeepLoc 2.0 predictions (29). Additional protein property annotations including probabilities for extracellular vesicle association were obtained from previously published datasets (30). Details are available in the **Supplemental Methods**.

## Results

### The circulating proteome differentiates leukemia immunophenotypes

To characterize the circulating blood proteome of pediatric acute leukemia patients at diagnosis, we performed high-throughput proteomics using the Olink Explore 3072 platform. The discovery cohort comprised of 87 serum samples: 75 from patients diagnosed with acute leukemia and 12 from healthy controls (**Supplemental Table S1**). After QC, two AML samples and one control were excluded, yielding a final dataset of 84 individuals: 73 patients diagnosed with leukemia (19 AML, 45 BCP-ALL, 9 T-ALL) and 11 healthy controls (**Table 1**; **Figure 1A**).

**Table 1.** Summary of patient characteristics (discovery cohort)

| **Characteristics** | **BCP-ALL**<br>**(n=45)** | **n** | **%** | **T-ALL**<br>**(n=9)** | **n** | **%** | **AML**<br>**(n=19)** | **n** | **%** |
| --- | --- | --- | --- | --- | --- | --- | --- | --- | --- |
| **Median age (range)** | 3 (1–14) |  |  | 13 (4–16) |  |  | 11 (0–17) |  |  |
| **Age group (years)** | <5 | 29 | 64.4 | <5 | 1 | 11.1 | <5 | 6 | 31.6 |
|  | 5–10 | 14 | 31.1 | 5–10 | 2 | 22.2 | 5–10 | 5 | 26.3 |
|  | 11–14 | 2 | 4.4 | 11–14 | 4 | 44.4 | 11–14 | 4 | 21.1 |
|  | ≥15 | 0 | 0 | ≥15 | 2 | 22.2 | ≥15 | 4 | 21.1 |
| **Sex** | Female | 22 | 48.9 | Female | 2 | 22.2 | Female | 10 | 52.6 |
|  | Male | 23 | 51.1 | Male | 7 | 77.8 | Male | 9 | 47.4 |
| **Year of diagnosis** | 2006 | 2 | 4.4 | 2007 | 2 | 22.2 | 2007 | 2 | 10.5 |
|  | 2007 | 4 | 8.9 | 2008 | 2 | 22.2 | 2008 | 2 | 10.5 |
|  | 2008 | 6 | 13.3 | 2010 | 1 | 11.1 | 2009 | 1 | 5.3 |
|  | 2009 | 5 | 11.1 | 2013 | 1 | 11.1 | 2010 | 7 | 36.8 |
|  | 2010 | 6 | 13.3 | 2014 | 1 | 11.1 | 2011 | 1 | 5.3 |
|  | 2011 | 7 | 15.6 | 2015 | 1 | 11.1 | 2012 | 2 | 10.5 |
|  | 2012 | 2 | 4.4 | 2017 | 1 | 11.1 | 2013 | 1 | 5.3 |
|  | 2013 | 4 | 8.9 |  |  |  | 2015 | 1 | 5.3 |
|  | 2014 | 3 | 6.7 |  |  |  | 2016 | 1 | 5.3 |
|  | 2015 | 4 | 8.9 |  |  |  | 2017 | 1 | 5.3 |
|  | 2016 | 2 | 4.4 |  |  |  |  |  |  |
| **White blood cell count** | **Median (range)** | 10.1<br>(1.4–109) |  | **Median (range)** | 264<br>(2.9–598) |  | **Median (range)** | 40.5<br>(1.6–495) |  |
|  | <50 | 44 | 97.8 | <50 | 3 | 33.3 | <50 | 11 | 57.9 |
|  | 50–100 | 0 | 0 | 50–100 | 0 | 0 | 50–100 | 5 | 26.3 |
|  | >100 | 1 | 2.2 | >100 | 6 | 66.7 | >100 | 3 | 15.8 |
| **% blasts in peripheral blood** | **Median (range)** | 33<br>(0–87) |  | **Median (range)** | 81<br>(1.4–96) |  | **Median (range)** | 38<br>(0–86) |  |
|  | <10 | 6 | 13.3 | <10 | 2 | 22.2 | <10 | 2 | 10.5 |
|  | 10–49 | 20 | 44.4 | 10–49 | 0 | 0 | 10–49 | 10 | 52.6 |
|  | 50–75 | 10 | 22.2 | 50–75 | 2 | 22.2 | 50–75 | 2 | 10.5 |
|  | >75 | 6 | 13.3 | >75 | 5 | 55.6 | >75 | 3 | 15.8 |
|  | Missing data | 3 | 6.7 |  |  |  | Missing data | 2 | 10.5 |
| **Subtype** | HeH | 20 | 44.4 |  |  |  | CEPBA | 1 | 5.3 |
|  | ETV6::RUNX1 | 25 | 55.6 |  |  |  | NPM1 | 1 | 5.3 |
|  |  |  |  |  |  |  | inv16 | 2 | 10.5 |
|  |  |  |  |  |  |  | 11q23 | 3 | 15.8 |
|  |  |  |  |  |  |  | t(8;21) | 2 | 10.5 |
|  |  |  |  |  |  |  | t(9;11) | 2 | 10.5 |
|  |  |  |  |  |  |  | Unknown | 8 | 42.1 |
| **Risk group** | Standard | 33 | 73.3 | Standard | 0 | 0 | Standard | 18 | 94.7 |
|  | Intermediate | 11 | 24.4 | Intermediate | 3 | 33.3 | High | 1 | 5.3 |
|  | High | 1 | 2.2 | High | 6 | 66.7 |  |  |  |
| **Relapse** | No relapse | 40 | 88.9 | No relapse | 7 | 77.8 | No relapse | 12 | 63.2 |
|  | Relapse | 5 | 11.1 | Relapse | 2 | 22.2 | Relapse | 7 | 36.8 |
| **Status** | Alive | 45 | 100 | Alive | 6 | 66.7 | Alive | 16 | 84.2 |
|  | Dead | 0 | 0 | Dead | 3 | 33.3 | Dead | 3 | 15.8 |

**Figure 1.**
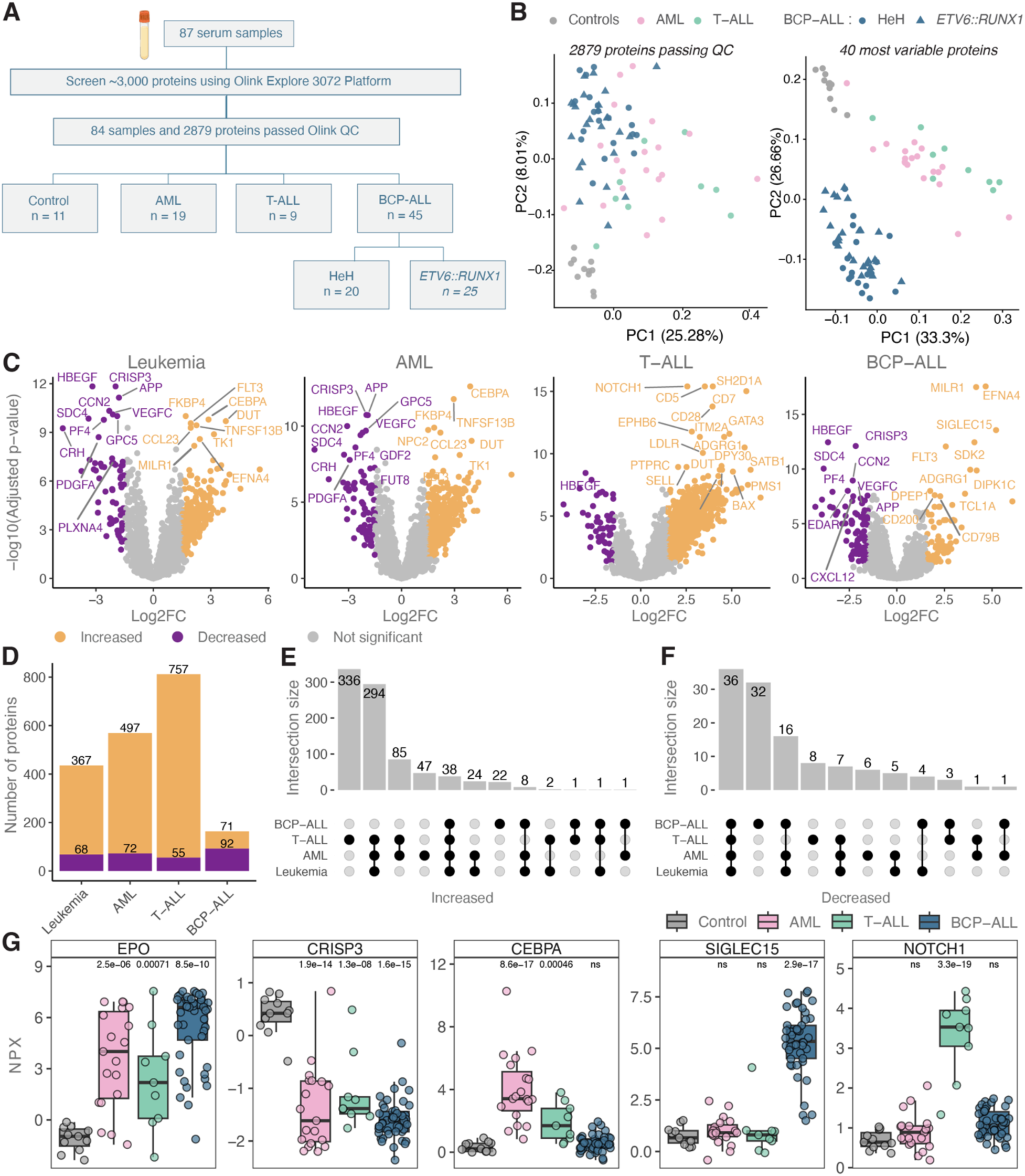
High-throughput serum proteomics reveals shared and subtype-specific protein expression patterns in pediatric acute leukemias. **(A)** Serum samples from healthy controls and pediatric patients diagnosed with acute myeloid leukemia (AML), T cell acute lymphoblastic leukemia (T-ALL), or B cell precursor ALL (BCP -ALL) were screened for the expression of >3,000 proteins using the Olink Explore platform. **(B)** Principal component analyses (PCA) of the proteins (n=2879) and samples (n=84) passing quality control (left) as well as the 10 most variable proteins per group (leukemia general, AML, T-ALL, and BCP-ALL; 40 proteins in total). Samples are colored according to the key above the plots. **(C-G)** Differential expression analyses were performed using *limma* with thresholds of |Log2FC| > 1.5 and FDR adjusted p < 0.05. First all leukemia samples were pooled and compared with controls then the three immunophenotypes (AML, T-ALL and BCP-ALL) were independently compared with controls **(C)** Volcano plots summarizing the analyses and highlighting proteins with increased (yellow) or decreased (purple) abundance in the different comparisons. Non-significant proteins are colored in grey and the twenty most significant proteins (based on lowest FDR-adjusted p-values) are labeled in each plot. **(D)** Stacked bar chart summarizing the number of increased (yellow) and decreased (purple) proteins identified in each comparison. **(E-F)** Upset plots showing the overlap of increased (E) and decreased (F) proteins across immunophenotypes. **(G)** Boxplots showing normalized protein expression (NPX) levels for selected proteins.

After QC, 2879 proteins remained (**Supplemental Table S2**) and unsupervised PCA revealed a clear separation between controls from leukemia samples but limited separation between immunophenotypes (**Figure 1B, left panel**). To explore the proteomic features that distinguished immunophenotypes, we identified the 10 most variable proteins per group (leukemia general, AML, T-ALL, and BCP-ALL; 40 proteins in total) and visualized their variance using PCA (**Figure 1B, right panel).** This supervised feature selection approach yielded improved visual separation between groups.

No clustering was observed by sex or patient age at diagnosis (**Supplemental Figure S1A-B**), suggesting that these variables did not drive the main variance in the dataset. Because samples were collected over an extended time period, we further assessed whether sample age (time in storage) influenced the serum proteomic profiles; however, no systematic clustering or trends were observed (**Supplemental Figure S1C**).

Next, we assessed whether circulating proteins could distinguish leukemia patients from controls; differential protein expression analysis revealed widespread alterations, most pronounced in T-ALL (n = 812 proteins), followed by AML (n = 569), and BCP-ALL (n = 163) (**Figure 1C-D and Supplemental Tables S4-7)**.

T-ALL also showed the largest set of unique proteins with increased abundance (n=336), followed by AML (n=47) and BCP-ALL (n=22) (**Supplemental Table S8-10**). A shared subset of 85 proteins was elevated in T-ALL and AML, while 38 proteins were elevated across all three immunophenotypes (**Figure 1E)**. Fewer proteins decreased overall, although BCP-ALL showed the largest set (n = 32), compared with T-ALL (n = 8) and AML (n = 6; **Figure 1F**).

Several biologically relevant proteins stood out (**Figure 1G**). Erythropoietin (EPO), a pleiotropic growth factor involved in erythropoiesis, bone remodeling, and innate immunity (31, 32), was consistently increased across all immunophenotypes. In contrast, CRISP3, a neutrophil-secreted extracellular matrix protein (33, 34), was uniformly reduced. These opposing signatures may reflect the bone marrow failure characteristic of leukemia and suggest a shared signature of impaired innate immunity. Distinct immunophenotype-specific protein expression profiles were also evident: CEBPA, a myeloid-associated protein (35), was exclusively elevated in AML; SIGLEC15, an adhesion molecule implicated in immune suppression (36) was exclusively elevated in BCP-ALL; and NOTCH1 as well as the early T-cell marker CD7 were exclusively elevated in T-ALL **(Figure 1G; Supplemental Figure S2)**

### Tracing the origins of differentially expressed blood proteins

To explore the cellular origins of altered circulating blood proteins in AML, BCP-ALL, and T-ALL, we first annotated each protein according to its subcellular localization: extracellular, intracellular, or membrane-associated (**Figure 2A**; **Supplemental Tables S11-13)**. Distinct distribution patterns emerged: proteins elevated in patients relative to controls were predominantly intracellular, whereas those less abundant were mainly extracellular. Membrane-associated proteins were more evenly represented among both over- and under-expressed groups (**Figure 2A**). Independent localization analyses revealed consistent patterns (**Supplemental Figure S3-4).**

**Figure 2.**
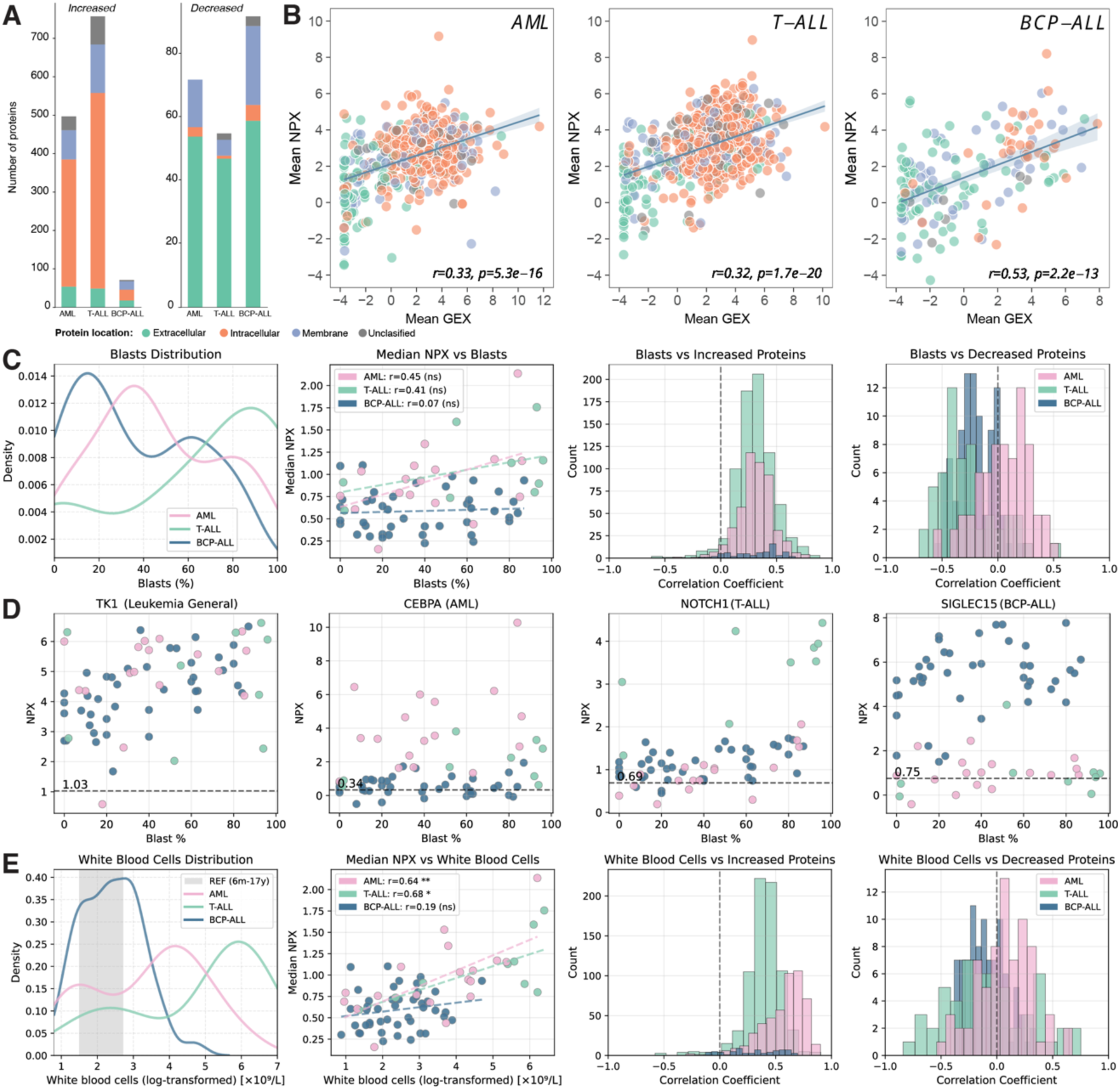
Protein annotations and clinical variables associated with serum protein abundance. **(A)** Stacked bar chart illustrating the number of proteins with increased or decreased abundance in each leukemia immunophenotype compared with healthy controls. Proteins are color-coded by UniProt cellular annotations; extracellular (green), intracellular (orange), membrane (blue), and unclassified (grey). **(B**) Correlation plots of average NPX values from Olink Discovery data (y-axis) versus corresponding mRNA expression data. Spearman’s correlation coefficients and p-values are noted in each panel. **(C-E)** Distributions and correlations of blast percentages (C-D) and log-transformed WBC counts (E) versus the median total protein expression by sample and immunophenotype. Density plots show blast and WBC distributions for AML (pink), T-ALL (green), and BCP-ALL (blue); a pediatric WBC normal reference range (6 months–17 years) is highlighted in gray. Scatter plots illustrate correlations between blast percentage or log-transformed WBC counts versus median NPX of all assayed proteins or selected proteins (TK1, CEBPA, SIGLEC15, and NOTCH; D). Pearson’s correlation coefficients and p-values noted. Bar charts summarize the distribution of correlation coefficients for blasts and WBC counts across significantly increased or decreased proteins. The number of altered proteins is indicated for each immunophenotype.

Because leukemic blasts circulate in the blood, we hypothesized that some of the membrane and intracellular proteins might reflect leukemia-cell origin. To test this, we compared serum protein abundance with transcript levels in leukemic blasts using RNA-seq data from diagnostic samples (BCP-ALL (HeH, n=46; *ETV6::RUNX1*, n=32), T-ALL (n=19), and AML (n=18) (**Figure 2B)** (27, 37). These samples largely derived from independent patients, and only few patients overlapped with the discovery cohort (3 HeH, 6 *ETV6::RUNX1*, and 2 T-ALL). Overall, correlations between mRNA and serum protein levels were modest (Spearman’s r=0.32–0.51), nonetheless, some gene–protein pairs showed strong concordance (e.g. DUT in all immunophenotypes, **Supplemental Tables S11-S13**). Other serum proteins such as EPO, had undetectable or low mRNA levels in blasts (**Supplemental Figure S5**), suggesting an origin from non-malignant cells.

To further assess the contribution of leukemic burden to serum protein levels, we examined associations between median NPX values across all proteins and peripheral blood counts at diagnosis. Percentage of peripheral blood blasts (PBB) showed modest correlation with median protein abundance in AML (Pearson’s r=0.452, p=0.069) and T-ALL (r=0.452, p=0.267), but not in BCP-ALL (r=0.069, p=0.665) (**Figure 2C**). Examination of NPX levels for top group-defining proteins revealed that even in samples with low (<20%) or no PBB, protein levels remained elevated, indicating their potential utility as diagnostic markers (**Figure 2D; Supplemental Figure S6**). In contrast, total WBC count correlated strongly with median protein abundance in T-ALL (r=0.679, p=0.044) and AML (r=0.639, p=0.003) (**Figure 2E**). In AML and T-ALL, which frequently present with elevated WBC counts (38), the strong correlations between protein abundance and WBC likely reflect the contribution of leukemic cell burden to circulating protein levels. Platelet and hemoglobin counts showed no association with overall protein abundance patterns (**Supplemental Figure S7**). In summary, although elevated WBC counts appear to drive changes in total serum protein abundance in leukemia samples, key putative disease marker proteins remain detectable irrespective of PBB or WBC levels, supporting their potential as putative biomarkers.

### Pathway and protein property analyses reveal shared and distinct signatures across acute leukemia immunophenotypes

Pathway enrichment analysis of the differentially expressed proteins identified 22 enriched pathways, encompassing both shared and immunophenotype-specific signatures (**Figure 3A; Supplemental Table S14-S15)**. Two pathways were consistently altered across all leukemia types: angiogenesis was decreased with reduced levels of canonical growth factors (e.g. VEGFC, PDGFA, PDGFB, HBEGF, CCN2, and ANGPT1), while E2F signaling was increased across all immunophenotypes with elevated levels of BCL2 and TK1 (**Figure 3B**).

**Figure 3.**
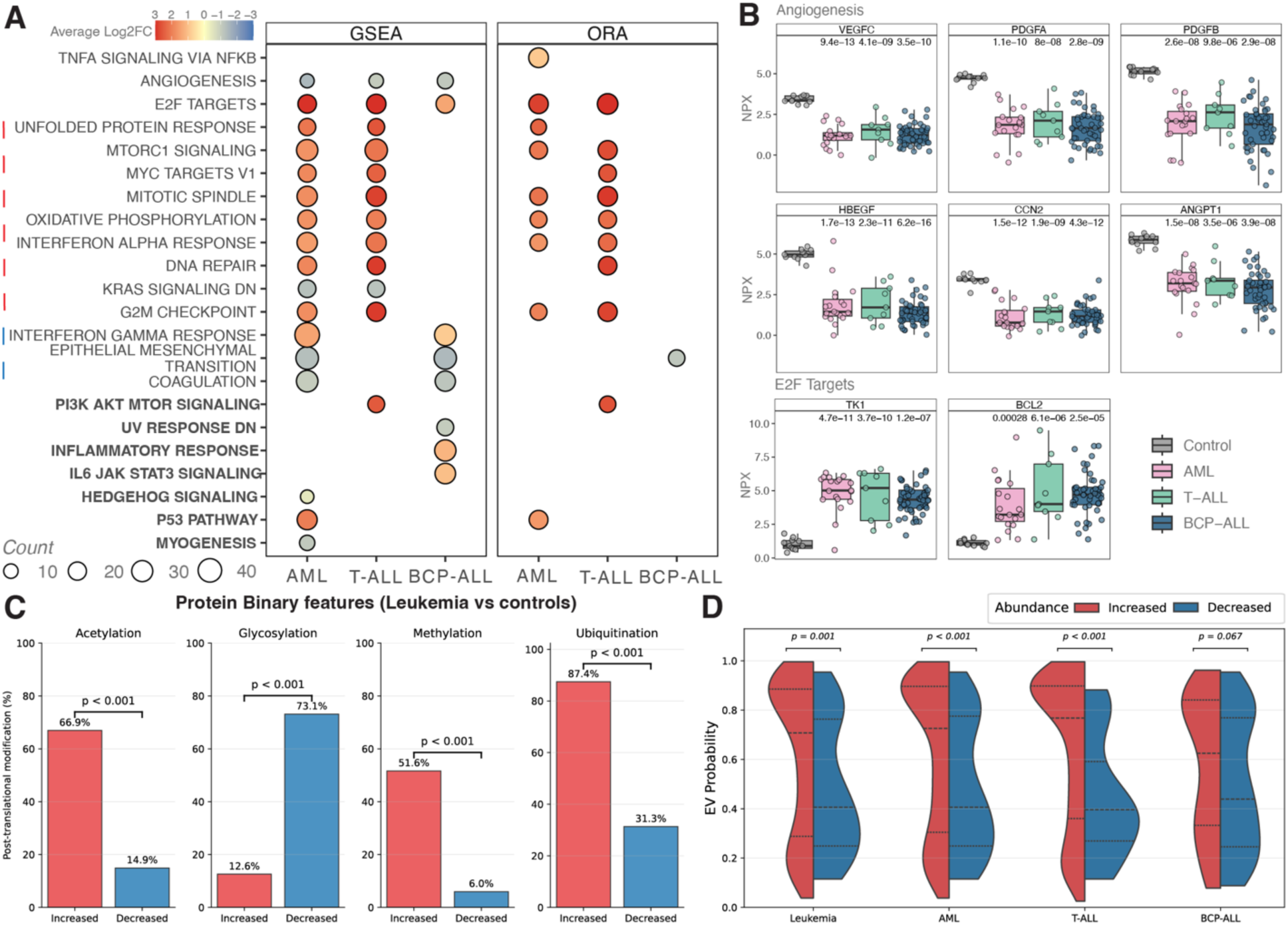
Pathway enrichment and protein properties analyses of dysregulated proteins reveals unique and distinct features in acute leukemia immunophenotypes. **(A)** Dot plot illustrates the overlapping and unique pathways identified between leukemia immunophenotypes (AML, T-ALL, or BCP-ALL) compared with healthy controls. The pathway analysis included a gene set enrichment analysis (GSEA) and an over representation analysis (ORA); the cutoff for determining statistical significance was FDR < 0,05. Each pathway is annotated with the number of proteins identified (count) and the average Log2FC of the proteins listed in the pathway to show if the pathways are up (red) or down (blue) regulated. **(B)** Boxplots illustrate the Normalized Protein Expression (NPX) levels of selected growth factors involved in angiogenesis and proteins involved with E2F targets in controls, AML, T-ALL, and BCP-ALL. Adjusted p-values are annotated above each group and represent the comparison made between each immunotype with healthy controls. **(C)** Split violin plots comparing probability of extracellular vesicle (EV) association distributions between proteins with increased versus decreased abundance. Statistical comparisons performed using Mann-Whitney U tests (**D)** Bar plots showing the percentage of proteins with specific post-translational modifications (acetylation, glycosylation, methylation, ubiquitination) among increased versus decreased proteins in leukemia patients. Statistical significance assessed using Fisher’s exact test. Protein properties for the human proteome and machine learning models for EV probability prediction used in this study are publicly available (67).

AML and T-ALL shared nine enriched pathways (**Figure 3A**, **dashed red line**), whereas AML and BCP-ALL only shared three (**Figure 3A, dashed blue line**), including interferon gamma response, epithelial-to-mesenchymal transition, and coagulation. Each immunophenotype also displayed unique signaling pathways, for example Hedgehog in AML, PI3K/AKT/mTOR in T-ALL, and IL-6/JAK/STAT3 in BCP-ALL (**Figure 3A, bold labels**).

Protein property analyses further revealed common features of highly abundant proteins across leukemia immunotypes, including solubility, hydrophobicity, and increased molecular weight (**Supplemental Figure S8**). Based on their annotated properties, proteins increased across all leukemia immunophenotypes were enriched for post-translational modifications such as acetylation, methylation, and ubiquitination while those decreased were more commonly annotated as glycosylated (**Figure 3C**) suggesting suppression of glycosylation-dependent secretory and immune functions (39). Distinct trends also appeared: proteins increased in AML and T-ALL (compared with controls) showed higher extracellular vesicle probability scores than the BCP-ALL group (**Figure 3D**). These results support the concept that leukemogenesis occurs and progresses in extracellular micro vesicle-enriched microenvironments(40), yet the differences in extracellular vesicle probability between the three immunophenotypes should be further evaluated.

### The genetic background of BCP-ALL impacts the circulating blood proteome

To investigate whether the genetic background of BCP-ALL influences the circulating blood proteome, we compared serum protein levels between the two most common molecular subtypes, HeH (n = 20) and *ETV6*::*RUNX1* (n = 25) (**Supplemental Table S16**). We identified 19 differentially expressed proteins, with ten increased in HeH and nine increased in *ETV6*::*RUNX1* (**Figure 4A**). Pathway enrichment revealed distinct signaling patterns in HeH, including interferon gamma response, IL-6/JAK/STAT3 signaling and KRAS signaling—aligning with the known enrichment of Ras-pathway mutations in this subtype (41–43) (**Figure 4B; Supplemental Tables S14–S14**). In contrast, coagulation was the only enriched pathway in *ETV6::RUNX1*. Effect size analysis of the 19 subtype-discriminating proteins showed a clear separation (**Figure 4C**). These 19 proteins also distinguished both subtypes from healthy controls and showed limited overlap with AML and T-ALL (**Supplemental Figure S9; Supplemental Table S19**).

**Figure 4.**
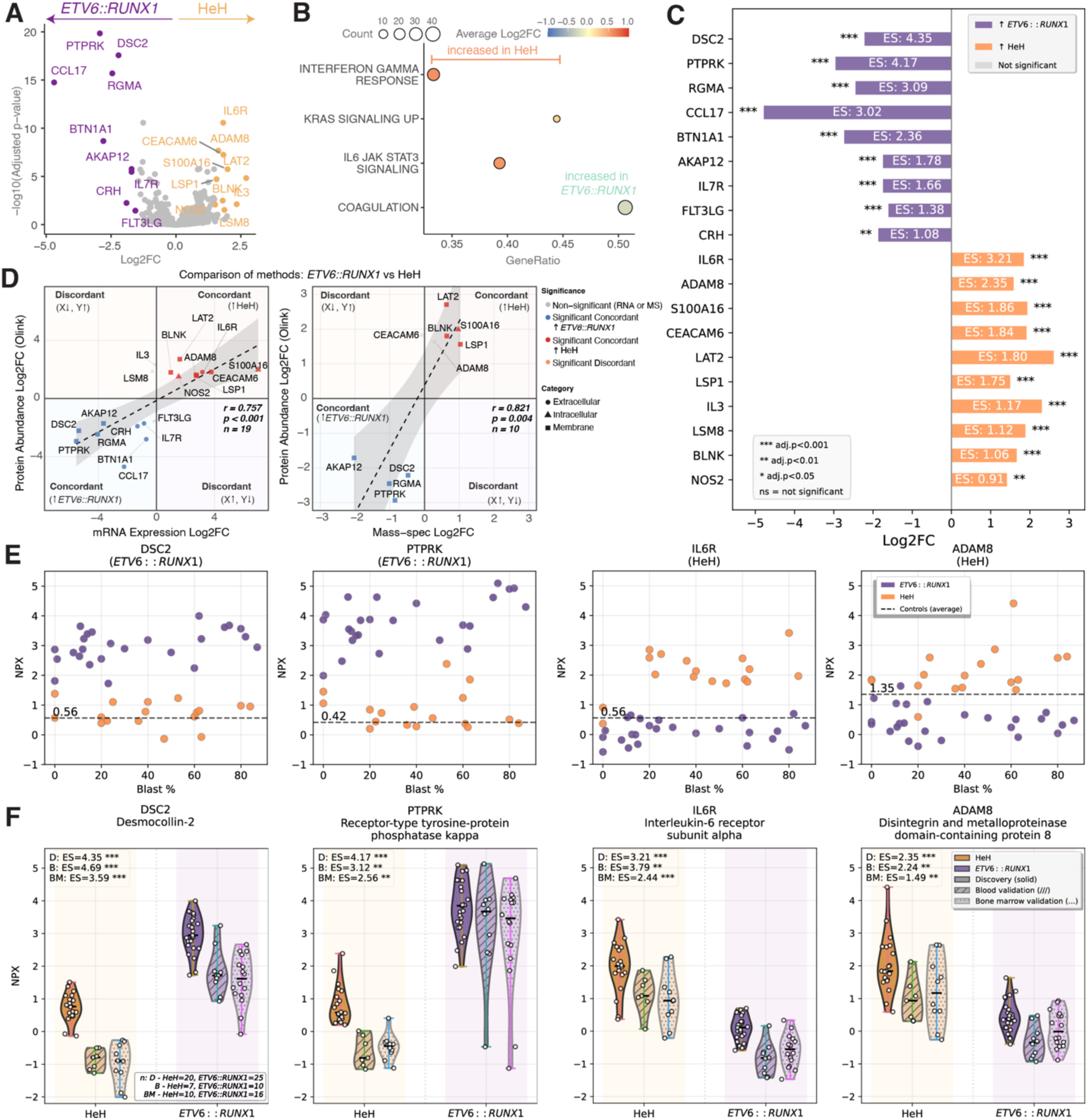
The genetic subtype of BCP-ALL influences the abundance of circulating blood proteins. **(A)** Volcano plot illustrates the comparison between HeH vs *ETV6::RUNX1.* Level of statistical significance was set at |Log2FC| ≥ 1.5 and FDR adjusted p-value < 0.05. Proteins increased in HeH are indicated in yellow and proteins increased in *ETV6*::*RUNX1* are purple. Non-significant proteins are colored in grey. **(B)** Dot plot illustrating the pathway enrichment based on the differentially expressed proteins (FDR < 0.05). Each pathway is annotated with the number of proteins identified (count) and the average Log2FC of all proteins in the corresponding pathway. **(C)** Forest plots show Log2 fold change values with effect sizes (ES) for each differentially expressed protein between HeH and *ETV6*::*RUNX1* samples in the discovery Olink data. Yellow bars represent proteins with higher levels in HeH samples, while purple bars indicate proteins with higher levels in *ETV6*::*RUNX1* samples. The ES significance is annotated for each protein; ***adj.p<0.001, **adj.p<0.01, *adj.p<0.05, and ns=not significant. **(D)** Quadrant plots comparing Log₂FC values of the 19 differentially abundant proteins identified between HeH and *ETV6*::*RUNX1* with the Olink Explore assay (y-axis) versus their corresponding gene expression levels from cellular RNA-seq data (x-axis, left plot) and cellular mass spectrometry data (x-axis, right). Points are colored by statistical significance (adjusted p < 0.05 in both methods) and concordance status with shapes indicate cellular compartment localization. Quadrants represent concordant higher abundance in HeH (upper right), concordant higher abundance *in ETV6*::*RUNX1* (lower left), and discordant regulation (upper left and lower right). Pearson correlation coefficients (r) with 95% confidence intervals and p-values are shown. Dashed lines represent linear regression with confidence bands. **(E)** Scatter plots showing the NPX levels of selected proteins compared with peripheral blood blast %. NPX levels of healthy controls were averaged and are indicated as dashed lines. **(F)** Violin plots of the top four discriminatory proteins across discovery and validation datasets. Plots show protein abundance distributions for Desmocollin-2 (DSC2), Receptor-type tyrosine-protein phosphatase kappa (PTPRK), and Interleukin-6 receptor subunit alpha (IL6R) and A disintegrin and metalloproteinase domain-containing protein 8 (ADAM8) in HeH (yellow) and *ETV6*::*RUNX1* (purple) samples. Data are shown for discovery samples (solid fill, n=45) as well as peripheral blood validation (diagonal hatching, n=17) and bone marrow validation (dotted pattern, n=26) samples. Individual patient values are shown as white dots with jitter. In the validation sets, 17 patients provided both blood and bone marrow plasma while 9 donors only provided bone marrow plasma. Sample sizes, effect sizes (ES), and statistical significance are indicated for each dataset. Black lines indicate medians.

Comparison with a MS–derived proteomics dataset (17) showed consistent patterns for 10 proteins (**Figure 4D, right**; **Supplemental Table S18**), indicating that several of the identified proteins reflect blast-intrinsic expression. Interestingly, the seven differentially expressed proteins annotated as extracellular were absent in the MS-based cellular dataset and showed minimal expression in RNA-seq data (log_2_(CPM)< 0.5; **Supplemental Table S18**) suggesting that those proteins unlikely derive from the leukemic blasts. Using publicly available scRNA-seq data from the Human Protein Atlas (HPA)(44) we checked for the potential cellular source of those seven extracellular proteins; this inspection revealed enrichment in different immune cells, for example *CCL17* in dendritic cells, *IL7R* and *FLT3LG* in T-cells, and *IL6R* in monocytes and neutrophils as well as secretory cells (**Supplemental Figure S10**).

We also examined the relationship between percentage of PBB and NPX levels for all 19 proteins (**Supplemental Figure S11**). Consistent with our previous observations, serum levels of the top subtype-discriminating proteins remained elevated irrespective of PBB status (**Figure 4E**).

Next, we validated the findings in an independent cohort profiled with the Olink HT platform (**Supplemental Table S19**). The 19 subtype-discriminating proteins were present in the Olink HT platform, all above LOD, and showed consistent directional changes across blood and bone marrow plasma (**Supplemental Figure S12; Supplemental Table S20)**. Among the proteins, DSC2, PTPRK, IL6R, and ADAM8 showed the largest effect sizes, highlighting them as strong subtype markers (**Figure 4F**). Collectively, these results suggest that the circulating proteome reflects the genetic background of BCP-ALL. We then revisited our ALL RNA-seq cohort to assess the specificity of the 19 subtype-discriminating proteins. We identified *ETV6::RUNX1* samples and compared the GEX values of genes coding for the *ETV6::RUNX1*-defining proteins to all other BCP-ALL subtypes; we did the same for the HeH (**Supplemental Figure S13**). This analysis revealed that *DSC2* has potential to distinguish *ETV6::RUNX1*, while *ADAM8* has potential to distinguish HeH from other BCP-ALL subtypes.

## Discussion

We characterized the circulating blood proteome of pediatric acute leukemia using the Olink Explore platform and minimally invasive blood samples. We profiled >3000 proteins and found that the circulating proteome captures similarities as well as immunophenotypic and genetic diversity in acute leukemia at diagnosis. These signatures extend beyond blast-intrinsic properties and reflect systemic processes, positioning circulating proteomics as a valuable complement to genomic and transcriptomic profiling in acute leukemia.

We observed elevated circulating BCL2 across immunophenotypes. Venetoclax, a selective BCL2 inhibitor, is emerging as therapy for pediatric leukemia (45), and high BCL2 expression has been associated with venetoclax response in other hematological malignancies (46). While the functional significance of circulating BCL2 remains to be determined, our results warrant further investigation into its potential as a biomarker for treatment stratification. More broadly, as immunotherapies become incorporated into pediatric leukemia treatment (10, 11), circulating proteins may provide new opportunities for real-time response monitoring (47).

Interestingly, our study revealed a larger number of dysregulated proteins in AML and T-ALL, than BCP-ALL. This may indicate similar leukemogenic programs of these two immunophenotypes. The observed differences may also partly reflect the strong positive correlation between WBC count and protein abundance, as both AML and T-ALL typically present with high WBC levels (38). However, WBC count alone is unlikely to fully account for all proteomic alterations, as some BCP-ALL patients also presented with high peripheral blast counts. These associations likely mirror the underlying biology, in which both circulating blasts and immunophenotype-specific mechanisms contribute to increased protein levels. Given that high WBC count is a hallmark of high-risk disease (48), some of these proteins could serve as biomarkers of disease severity or leukemic cell burden and may also provide insight into mechanisms driving excessive leukocyte proliferation. Notably, several leukemia-associated proteins remained detectable even in patients with low or absent PBB. This observation likely reflects a combination of direct and indirect mechanisms. Some detected proteins may originate from leukemic cells despite low circulating blast percentages, as the sensitivity of the proteomic assay allows for detection of less abundant proteins. For example, T-cell surface markers CD5 and CD7 (49, 50) were found to be elevated in T-ALL samples, including in samples were PBB was low. In addition, a subset of proteins may derive from non-leukemic cells and reflect systemic responses to leukemia, such as immune activation, inflammatory signaling, or disrupted hematopoiesis, such as EPO in which renal origin is well established (51). Such indirect effects may persist even when leukemic blasts are largely confined to the bone marrow. Nevertheless, this finding is important, as the detection of blasts in peripheral blood is typically a key indicator of leukemia and prompts further examinations, including bone marrow analysis. Conversely, a low number of PBB may delay diagnosis. Moreover, the morphological identification of blasts is both challenging and subjective, complicating the initial subclassification of acute leukemias prior to comprehensive bone marrow analysis (52, 53). The detection of immunophenotype- and subtype-specific blood proteins therefore might offer a valuable complementary approach for early detection of leukemia, as well as for supporting molecular classification and disease monitoring, particularly in cases lacking circulating blasts.

Although many of the identified proteins likely originate from blasts, several point to a broader microenvironmental remodeling. For instance, while leukemic cells are known to promote angiogenesis in the bone marrow (54), we observed a reduction in angiogenic proteins across immunophenotypes, suggesting changes in the peripheral vasculature. Vascular complications such as hypertension and cardiac impairment are well-documented side effects of anti-leukemic therapies (55), but our findings raise the possibility that the disease itself may contribute to vascular vulnerability, potentially predisposing patients to these complications (56). We also noted elevated EPO across all immunophenotypes, likely reflecting compensatory responses to anemia and bone marrow dysfunction. Given EPO’s additional role in vascular and immunomodulatory effects (57), this observation further underscores how systemic physiological processes can be reflected in the circulating proteome.

Strikingly, we found that circulating proteins differed between BCP-ALL patients in the HeH and *ETV6*::*RUNX1* subtypes. These findings were validated in an external Olink dataset. Established subtype-defining markers of HeH (S100A16, BLNK) and *ETV6*::*RUNX1* (DSC2, RGMA) (17, 27, 58–60) were among the top validated proteins. Beyond proteins of interest for diagnostic subtyping, we identified proteins of clinical interest. CCL17, a chemokine elevated in *ETV6*::*RUNX1*-positive patients, has previously been shown to normalize after treatment (61), suggesting potential utility as a treatment-response biomarker. In the HPA scRNA-seq database, *CCL17* is mainly expressed by dendritic cells (62) raising the possibility of systemic, subtype-specific immune modulation that warrants further investigation. Importantly, most subtype-discriminating proteins were not differentially abundant in AML or T-ALL (compared to controls). This is clinically relevant, as pediatric patients with leukemia frequently present with overlapping symptoms. However, these comparisons are presented as observations rather than evidence of biomarker specificity, and extensive pediatric reference data will be needed to substantiate this.

This study has limitations. The number of samples per immunophenotype was modest, only two genetic subtypes of BCP-ALL were investigated, and only diagnostic samples were analyzed. However, we validated our observations using an external Olink dataset, for HeH and *ETV6*::*RUNX*1-positive cases. Given the well-described molecular differences between these subtypes at the epigenomic, transcriptomic, and cellular protein levels (63), we leveraged existing knowledge to demonstrate the likely origins of subtype-defining proteins. However, direct sample-level correlations could not be investigated because the transcriptomic and MS proteomic datasets largely derived from independent cohorts and only few patients overlapped with the discovery set. While our discovery cohort included only serum samples, the external dataset consisted of plasma from both blood and bone marrow, reaffirming the robustness of the Olink PEA method and our observations. Future studies incorporating serial and tissue-paired samples as well as additional subtypes will be critical to define more precisely the biological and clinical significance of the circulating proteins identified here.

In conclusion, the circulating proteome reflects both immunophenotypic and genetic diversity in pediatric leukemia, encompassing tumor-intrinsic programs as well as systemic physiological responses. Our study provides proof of concept that circulating proteins contain a rich layer of information worth exploring. This is particularly relevant given the increasing use of immunotherapies (64), and need for scalable, less complex approaches for diagnosis and disease monitoring (65). Blood proteomics offers distinct advantages such as minimally invasive sampling (particularly suitable for pediatric patients) and feasibility for repeated measurements (66). While our findings are exploratory, they highlight the potential of proteins as informative biomarkers and lay the foundation for future studies aimed at translating blood proteomics into clinical research and, ultimately, practice.

## Supporting information

Supplemental Methods, Figures, and References

## Data and code availability

The Olink proteomics data generated in this study will be deposited in the SciLifeLab Data Repository upon publication. The data is available under restricted access due to data privacy regulations aimed at protecting sensitive personal information. All other data supporting the findings of this study are available within the supplemental material. The GEX data for ALL are available at GEO under accession number GSE227832 and for AML under controlled access via https://doi.org/10.17044/scilifelab.13105229.v1 (https://figshare.scilifelab.se/). Codes used to generate figures in this study are deposited in Github (https://github.com/Molmed/BloodProteomicsALL).

## Acknowledgements

This study was supported by the Erik, Karin and Gösta Selander’s foundation (to APE), the Swedish Cancer Society #22-2395 (to JN), Swedish Childhood Cancer Fund #TJ2020-0039 (to AH/APE) and #HFT2023-0011 (to JN), the Swedish Association for Medical Research, SSMF #PG-23-0420-H-01 (to MAG), National Funding for Research and Education in Swedish Health Care Regions (ALF; to AH), Jane and Aatos Erkko Foundation and Sigrid Juselius Foundation (to OL and MH), the Foundation for Tampere University Hospital (to OL), the Foundation for Pediatric Research (to OL), the Swedish Research Council #2024-02251 under the frame of EP PerMed (GEPARD-2; to JN, OL), and the Finnish Ministry of Education (to TT).

We thank the SciLifeLab National Genomics Infrastructure (NGI) and Affinity Proteomics Uppsala for assisting with data generation. NGI is funded by SciLifeLab, the Knut and Alice Wallenberg Foundation, and the Swedish Research Council. We thank the NOPHO Leukemia Biobank for providing access to the serum samples used in this study. We thank Emil Sundberg for providing the control samples as well as Henrik Gezelius and Jonne Nieminen for technical support in the lab. We sincerely thank the patients and their families for their participation and contribution to this research.

## Author contributions

Conceptualization: **APE, MAG, AH, JN;** Investigation: **APE, LO;** Data curation: **APE, MAG, DG, TT, OK, AL, MLW;** Formal analysis: **APE, MAG, DG, TT, OK, AL, MLW, CH, MÅ;** Resources: **JP, AH;** Writing – original draft: **APE, MAG, JN;** Writing – review & editing: **DG, TT, OK, AL, LO, MH, OL, CH, SM, LH, MÅ, JP, AH, MLW;** Visualization: **APE, MAG, DG, OK, AL;** Supervision: **AH, JN;** Project administration: **JN;** Funding acquisition: **APE, MAG, MH, OL, AH, JN.**

## Notes

### Competing Interest Statement

The authors have declared no competing interest.

### Author Declarations

The study (discovery cohort) was conducted in accordance with the Declaration of Helsinki and approved by the Swedish Ethical Review Authority [#2021-05213]. Samples from the validation cohort were collected under the ethical permit #R13109 and approved by the Pirkanmaa Hospital District Ethical Committee in Finland.

