## Supplemental Methods, Figures, and References for "The circulating blood proteome of childhood acute leukemia"

Serum protein profiling (discovery cohort)

Proteomic profiling was performed using the Olink Explore 3072 platform, a highly-multiplexed, next-generation sequencing-based extension of the Proximity Extension Assay which employs matched antibody pairs conjugated to oligonucleotides that bind to target proteins, hybridize and extend to form a unique DNA barcode, which is subsequently amplified and quantified. The Explore 3072 library consists of eight 384-plex panels with internal controls and ~3000 target proteins related to well-established biomarkers and exploratory proteins within diverse biological processes such as cardiometabolic, inflammation, neurology, and oncology.

Sample preparation and sequencing (Illumina NovaSeq 6000, SP flow cell, 100-cycle kit v1.5) were performed according to the manufacturer's protocol. Sequencing data were processed using Olink Explore software (version 3.6.1) which generate Normalized Protein eXpression (NPX) values (relative, log2-transformed units derived after normalization to internal controls and a correction factor). NPX values enable relative comparison of protein levels across samples within the same study, but do not represent absolute protein concentrations (1).

Processing of publicly available RNA-seq data

The ALL RNA-seq dataset was aligned to the hg38 reference genome, while the AML RNA-seq dataset was aligned to hg19. For the ALL dataset, batch-correction was performed using CombatSeq (2) to correct for technical biases caused by the different library preparation methods (TruSeq, RNA Access, and Script-Seq). The AML RNA-seq dataset was not subjected to batch correction as all samples were prepared with the Illumina TruSeq Stranded total RNA (ribosomal depletion) library (3). The raw count matrices were normalized with Gene Length corrected trimmed mean of M-values (GeTMM)(4) to adjust for both gene length and library size. The resulting counts were log2 transformed. We harmonized the ALL and AML gene expression datasets to a single gene annotation (Ensembl GRCh38, version 113), resulting in the same set and number of genes (60 048) in both. The processed datasets were merged into a single expression matrix comprising 135 samples.

Plasma protein profiling (validation cohort)

Plasma samples were collected during mononuclear cell isolation, after the density gradient centrifugation for which blood or bone marrow samples were diluted 1:1 with HBSS (Gibco). Sequencing was performed on an Illumina NovoSeq 6000 sequencer at according to the manufacturer's protocol. Sequencing data were processed with Olink Explore software (version 6.7.2 and version 7.4.7) to generate NPX values. Fixed LOD values were downloaded from https://olink.com/knowledge/documents and integrated into the data. Statistical significance of protein abundance differences between subtypes was assessed using the Mann-Whitney U test (two-sided) for each protein, with false discovery rate correction using the Benjamini-Hochberg method to account for multiple testing. Effect sizes were calculated using Cohen's d. To remove technical batch effects, we applied ComBat harmonization using the *neuroCombat* Python implementation on the protein expression matrix with batch as a covariate.

Protein property annotations

To investigate the relationship between protein physicochemical properties and differential abundance in leukemia, we calculated Spearman correlations between continuous protein features and Log2 FC values. For differentially abundant proteins in AML, BCP-ALL, T-ALL, and Leukemia (compared to controls), correlations were computed between protein Log2FC values and physicochemical properties with continuous values (e.g. molecular weight, hydrophobicity, surface accessibility measures, secondary structure components, aggregation propensities). Multiple testing correction was applied (Benjamini-Hochberg method) across all tested correlations. To examine specific protein features that showed significant correlations with differential abundance, we performed detailed comparisons between increased and decreased proteins for selected continuous and categorical variables. For continuous features, we applied Mann-Whitney U tests to assess statistical differences between increased and decreased protein groups. For categorical features representing post-translational modifications (acetylation, glycosylation, methylation, and ubiquitination), we calculated the percentage of proteins known to have each modification within increased and decreased groups and used Fisher's exact tests to evaluate statistical significance of the differences.

Protein subcellular annotations

Extended subcellular localization predictions were generated using the deep learning-based multi-label predictor DeepLoc 2.0 (5) which identifies protein localization across ten cellular compartments: nucleus, cytoplasm, extracellular space, mitochondrion, cell membrane, endoplasmic reticulum, Golgi apparatus, lysosome/vacuole, peroxisome, and plastid. Only statistically significant proteins (FDR adjusted p-value $< 0.05$ and Log2 fold change ≥1.5) were included in this analysis.

Functional enrichment analysis of dysregulated proteins in leukemia.

The Search Tool for the Retrieval of Interacting Genes/Proteins-database (STRING-DB) was used to annotate differentially expressed proteins (adjusted p-value $< 0.05$; Log2 fold change ≥1.5) to tissue origin and subcellular compartment. Analysis was performed using the proteins in the Olink Explore 3k panel as background.

### Supplemental Figures

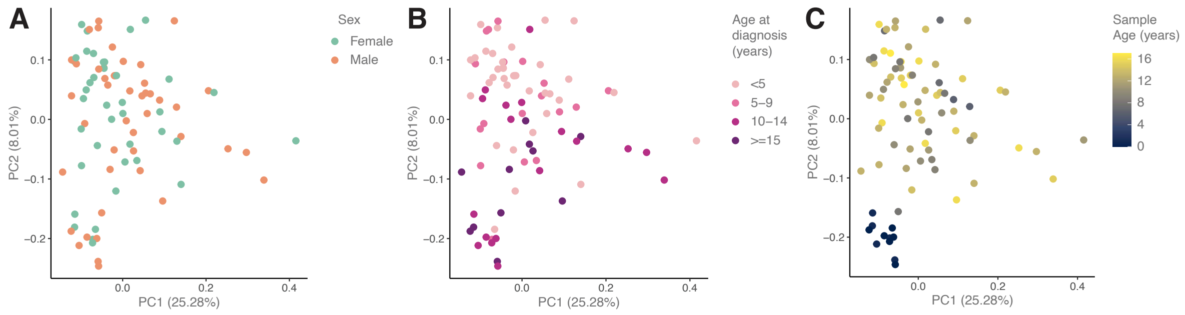


**Supplemental Figure S1. No clear separation based on sex, age at diagnosis or sample age in the discovery cohort.** Principal component analyses (PCA) of all serum samples in the discovery cohort colored by **(A)** sex **(B)** patient age at diagnosis and **(C)** sample age. Samples were assayed on the Olink Explore 3072 platform and the 2879 proteins passing quality control were used for the PCA.

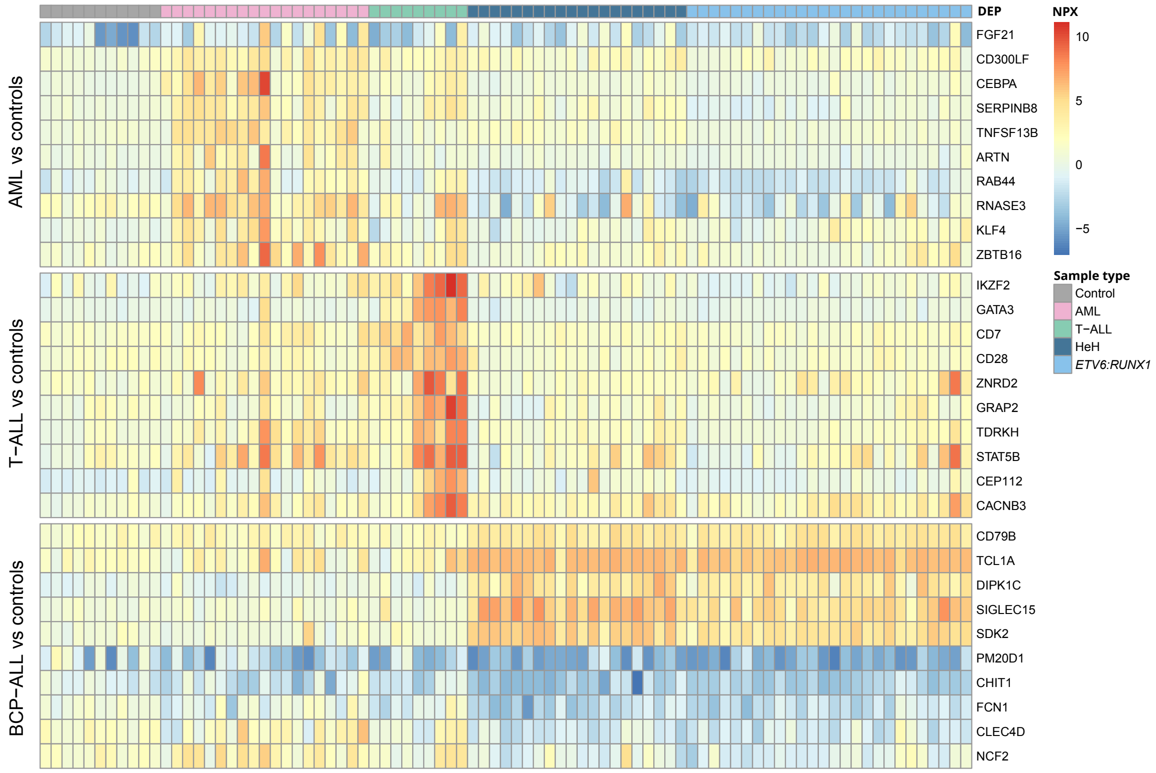


**Supplemental Figure S2. Serum protein expression differentiates immunophenotypes in pediatric acute leukemia.** Heatmap showing the normalized protein expression (NPX) value of the top ten unique differentially expressed proteins (right y-axis) identified in each immunophenotype when compared to controls. Leukemia patients (top x-axis) are labeled by immunophenotype or subtype (BCP-ALL) and controls are in grey. Differential expression was assed using *limma* (FDR-adjusted p-value < 0.05 and |Log2FC| > 1.5).

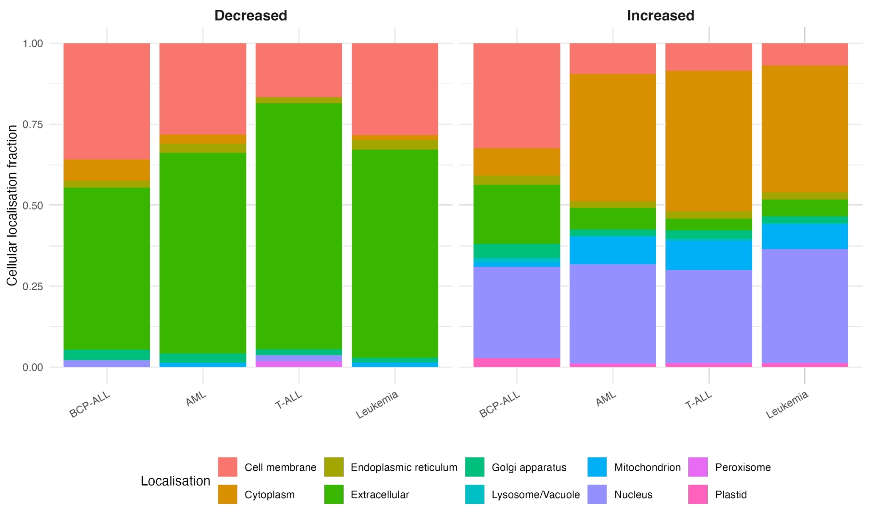


**Supplemental Figure S3. Subcellular localization patterns of differentially expressed proteins across leukemia subtypes.** Stacked bar plots show the proportion of proteins predicted to localize in different cellular compartments for proteins with decreased (left panel) and increased (right panel) abundance levels. Proteins were classified as increased or decreased based on statistical significance (adjusted p-value < 0.05 and absolute Log2 fold change ≥1.5) compared to controls. Subcellular localization predictions were obtained using DeepLoc 2.0, a deep learning-based multi-label predictor that identifies protein localization across 10 cellular compartments: nucleus, cytoplasm, extracellular space, mitochondrion, cell membrane, endoplasmic reticulum, Golgi apparatus, lysosome/vacuole, peroxisome, and plastid.

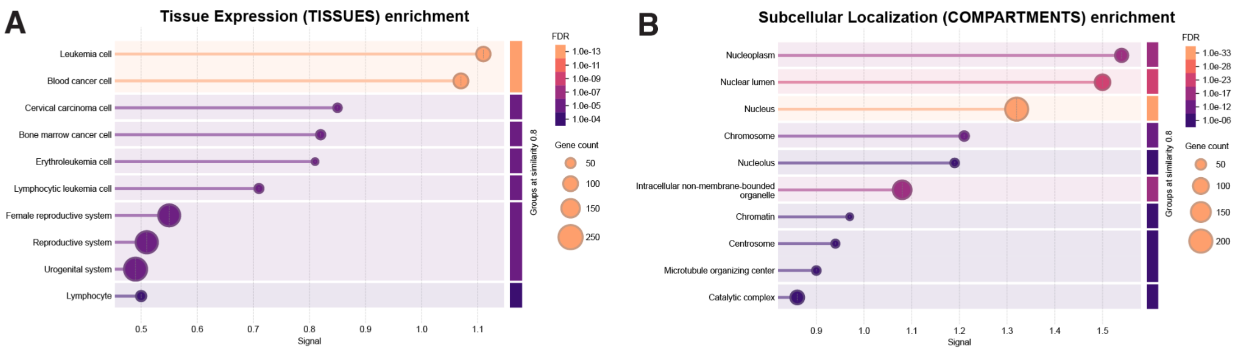


**Supplemental Figure S4. Functional enrichment analysis of dysregulated proteins in leukemia.** STRING-DB analysis of **(A)** tissue and **(B)** subcellular compartment enrichment analysis of proteins with significantly higher abundance in leukemia patients versus controls (adjusted p-value < 0.05; Log2 fold change ≥1.5). Dot size represents gene count; color intensity indicates FDR-adjusted p-value. Analysis performed using the proteins in the Olink Explore panel as background.

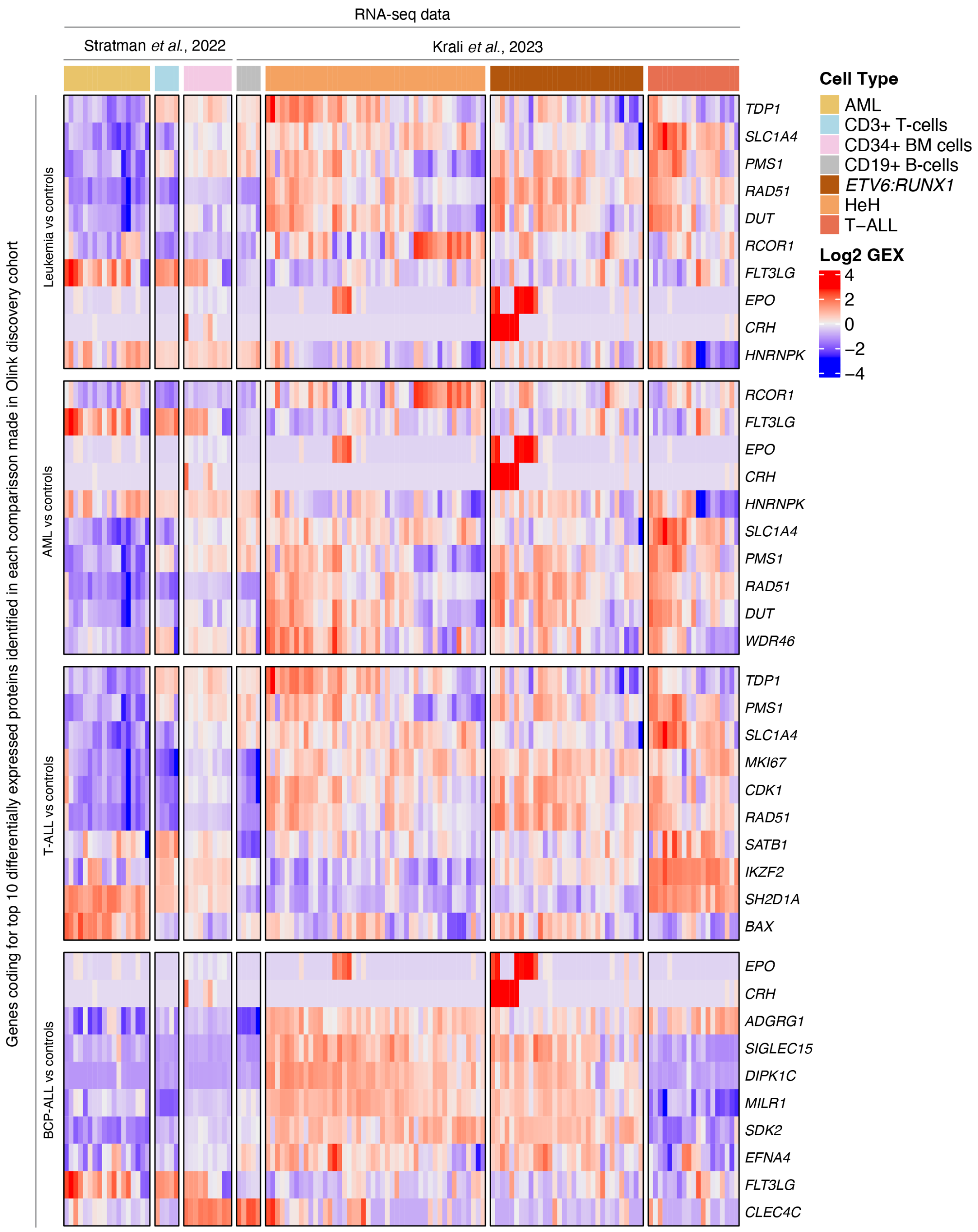


**Supplemental Figure S5. Gene expression profile of leukemic blasts, B-cells, T-cells, and bone marrow cells**. Heatmap of genes coding for the top ten differentially expressed proteins identified in the Olink discovery dataset. Publicly available bulk RNA-sequencing data of patient-derived AML, BCP-ALL, and T-ALL blasts were used as well as CD19+ B-cells, CD3+ T-cells and CD34+ bone marrow (BM) cells. BCP-ALL samples are annotated as *ETV6::RUNX1* or HeH. Gene expression data were obtained from Stratman *et al.* 2022 and Krali *et al.*, 2023 (accession number GSE227832).

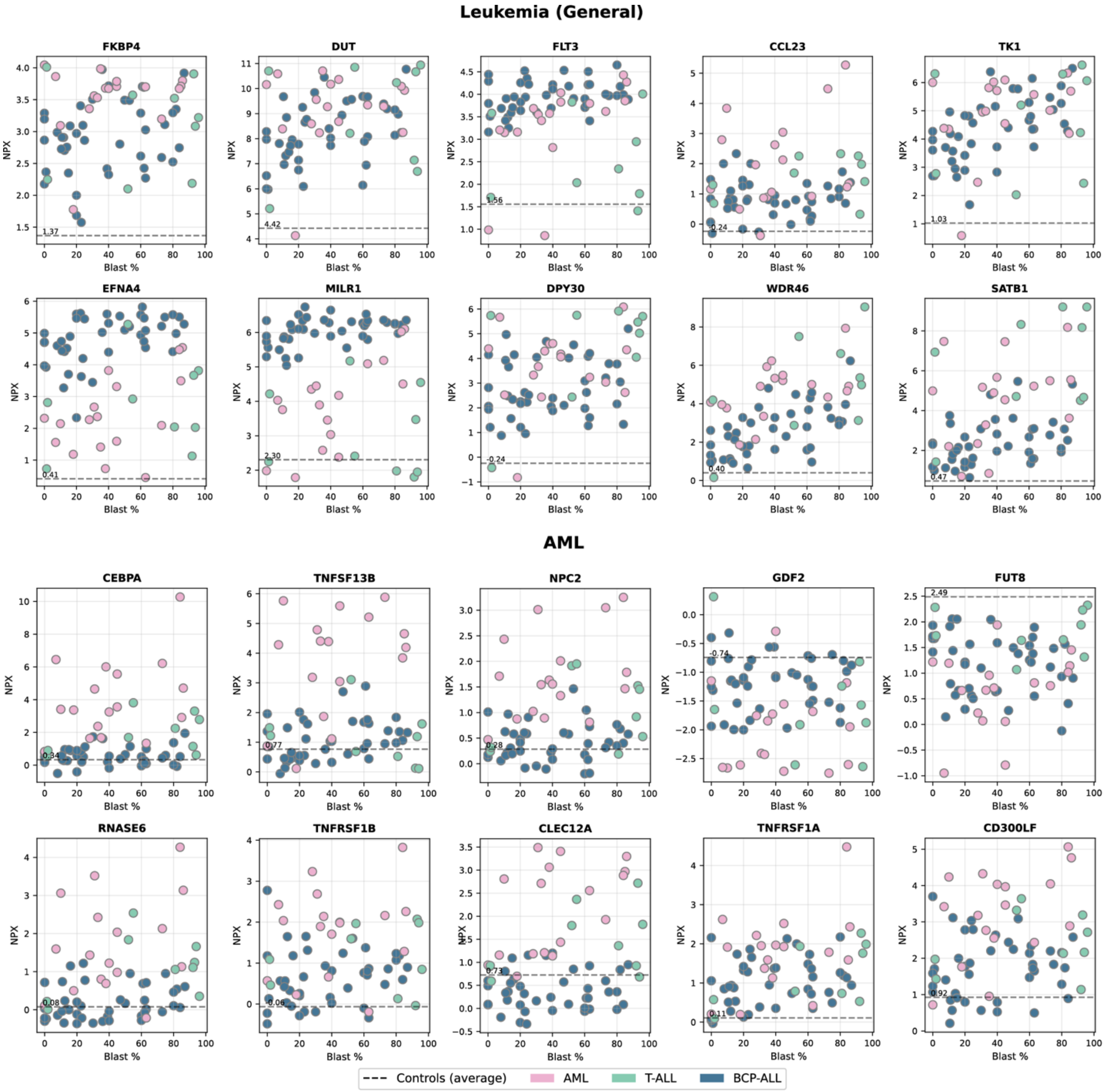


**Supplemental Figure S6. Protein abundance levels remain high in patients' leukemia patients with low or no peripheral blood blasts (PBB).** *Figure and legend continued on next page*

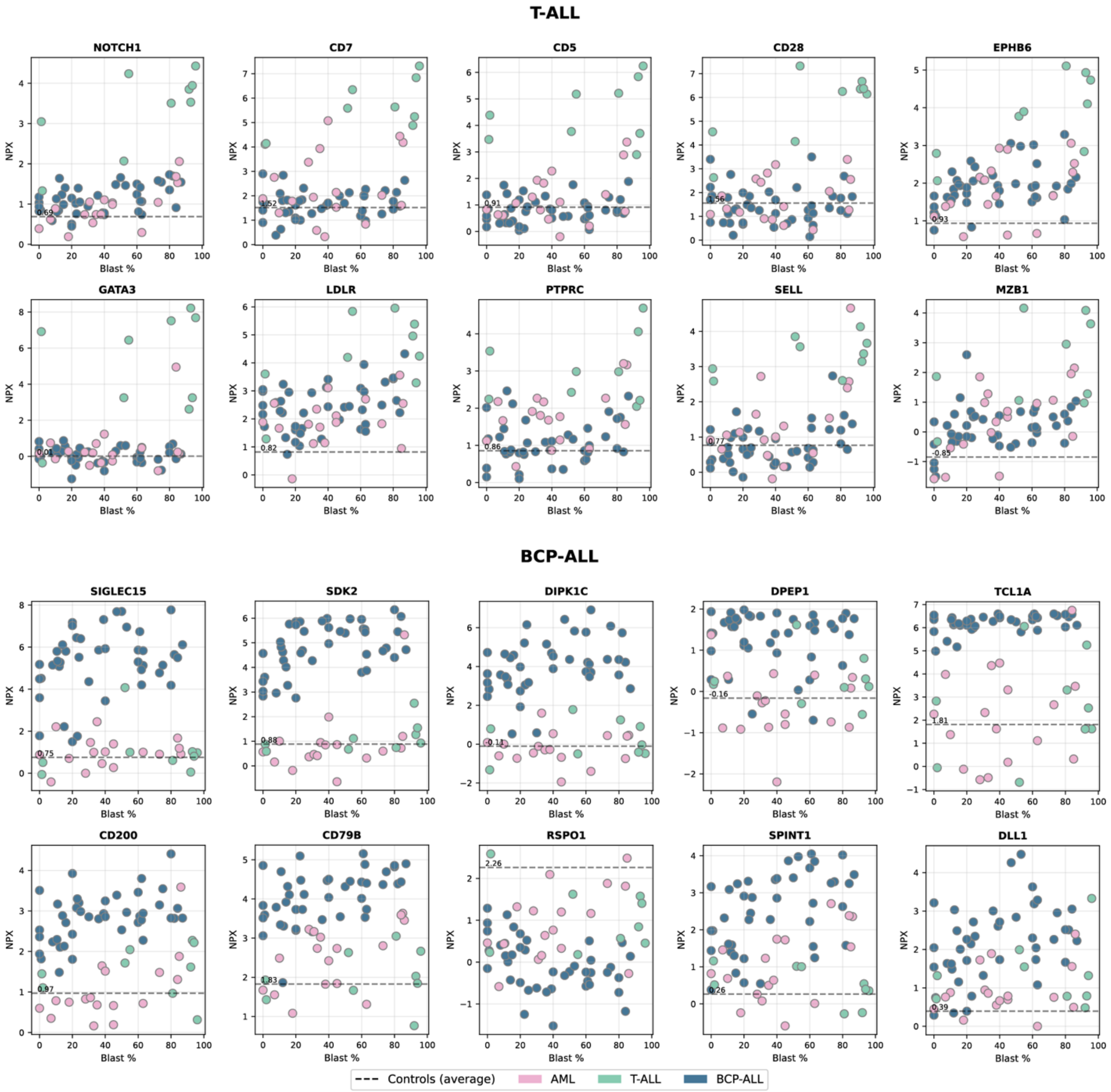


**Supplemental Figure S6-*continued*.** Scatter plots showing NPX values (y-axis) of selected proteins in all leukemia patients compared with PBB (x-axis). Patient samples are colored by immunophenotype; AML (pink) T-ALL (green) and BCP-ALL. NPX levels of control samples were averaged and are indicated by a dashed line.

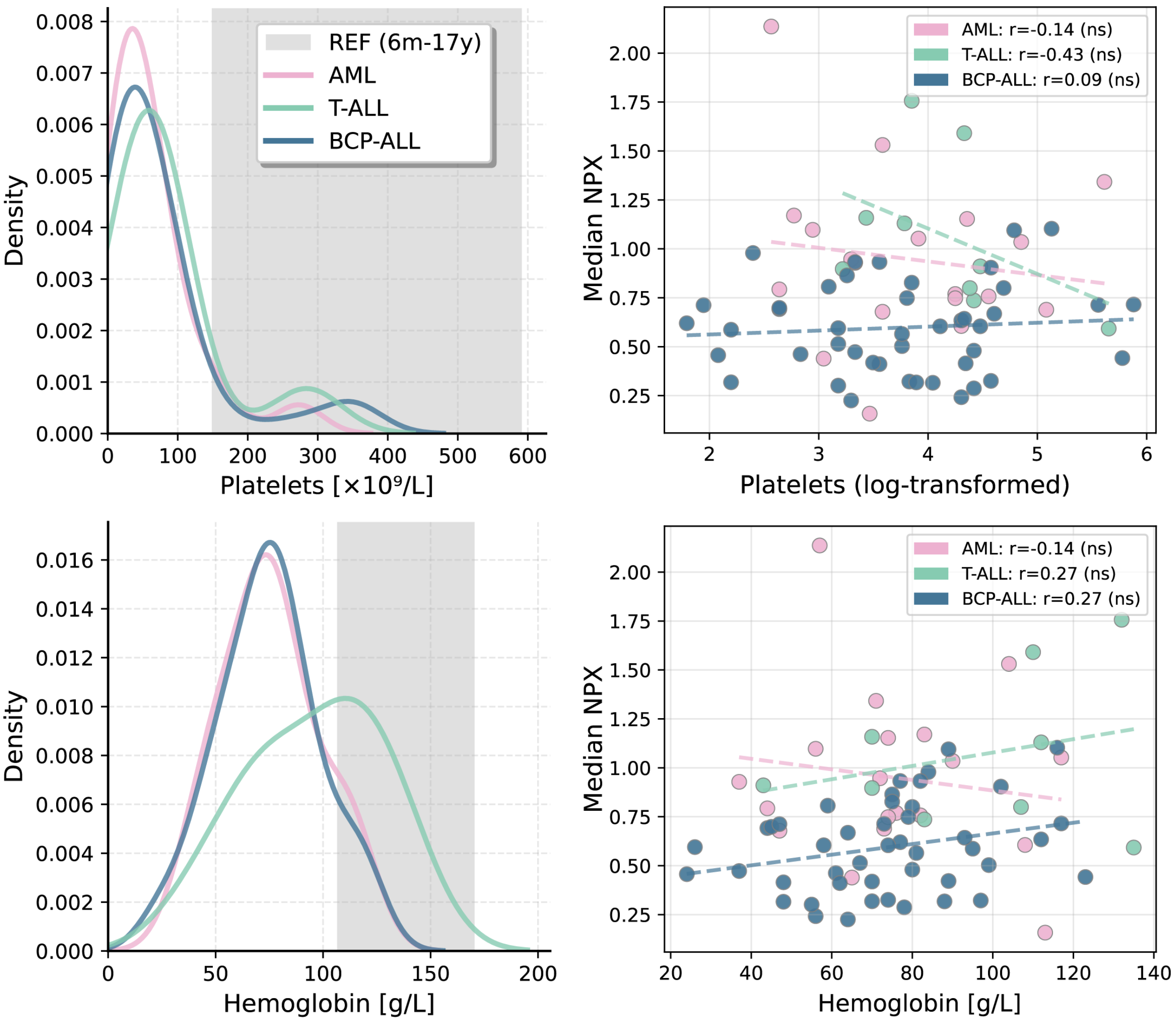


**Supplemental Figure S7. Platelet and hemoglobin distributions with median protein expression correlations across leukemia immunophenotypes.** The line histograms (left) illustrate the density distributions of platelet counts (top) and hemoglobin counts (bottom) for leukemia patients included in the discovery cohort. Pediatric reference ranges (6 months-17 years) are gray. The scatter plots (right) display the correlation of log transformed platelet counts (top) and hemoglobin (g/L, bottom) with median normalized protein expression (NPX) for every leukemia patient across immunophenotypes. Pearson correlation coefficients and p-values are indicated on the plots. Immunophenotypes are color-coding as following: AML (pink), T-ALL (green), and BCP-ALL (blue).

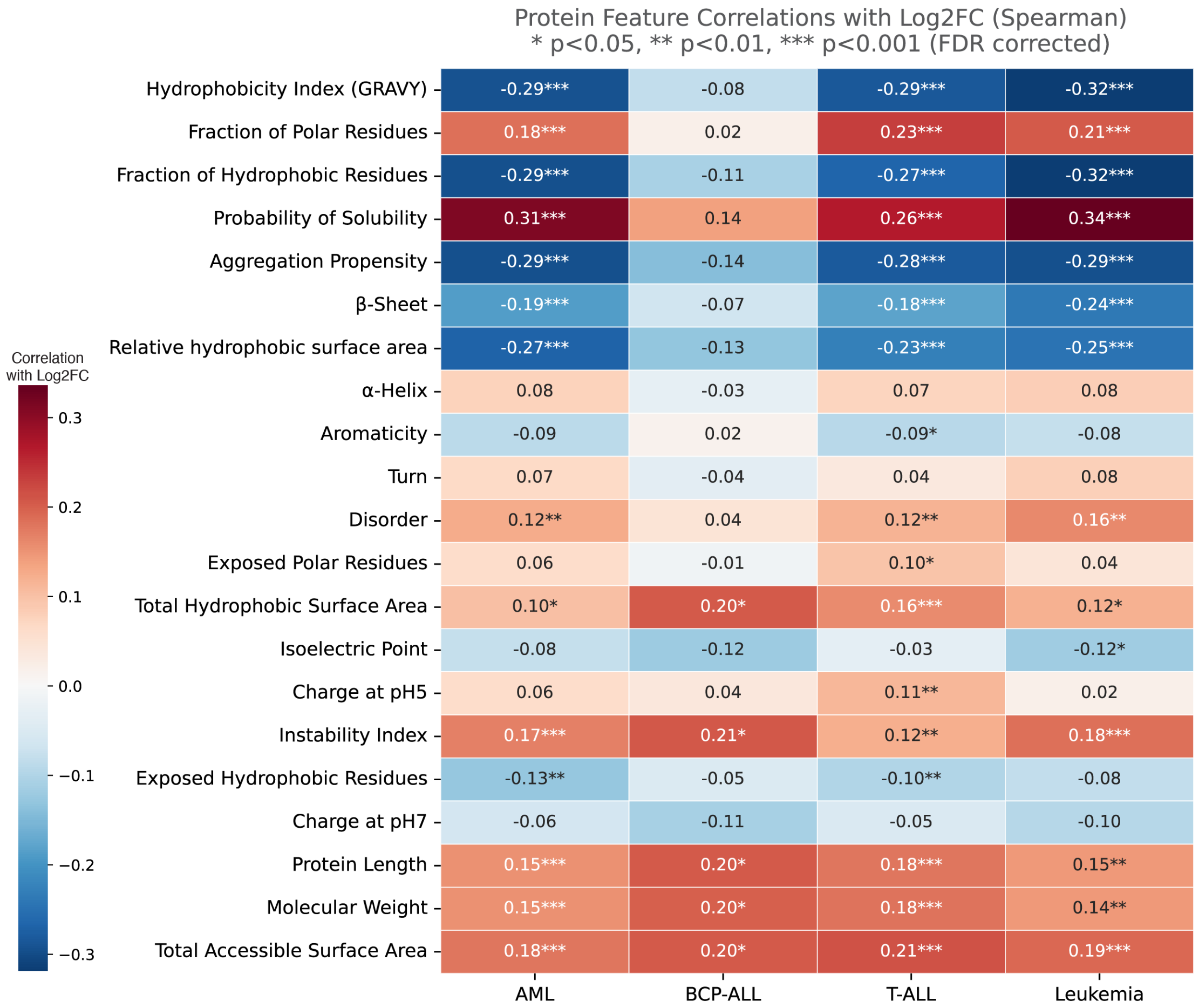


**Supplemental Figure S8**. **Protein Features correlations**. Heatmap illustrates Spearman correlations between protein physicochemical properties and Log2 fold changes across differentially abundant proteins (AML, BCP-ALL, T-ALL, and Leukemia compared with controls). Color intensity represents correlation strength; asterisks indicate statistical significance after FDR correction.

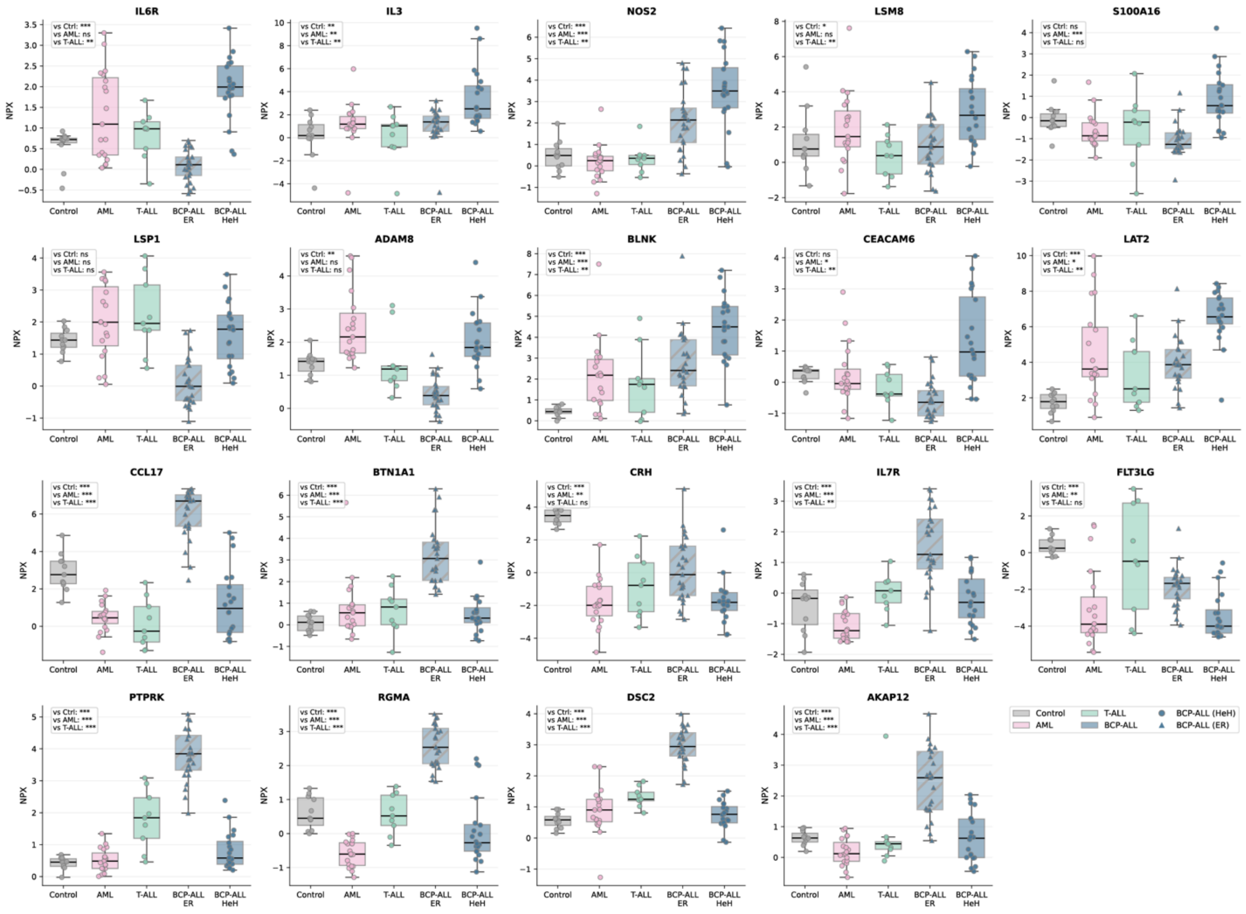


**Supplemental Figure S9.** Boxplots illustrate the normalized protein expression (NPX; y-axis) of the 19 subtype-defining proteins across all leukemia patients and controls (x-axis). Protein expression (NPX) in the indicated BCP-ALL subtype (*ETV6::RUNX1* or HeH) and Control, AML, and T-ALL groups was compared using two-sided Mann–Whitney U tests. For each protein, p-values were computed for the three predefined comparisons (subtype vs Control/AML/T-ALL). P-values were adjusted using the Benjamini–Hochberg false discovery rate (FDR) correction. Statistical significance is indicated by stars based on adjusted p-values: ns, p < 0.05 *, *p < 0.01 **, p < 0.001 ****.

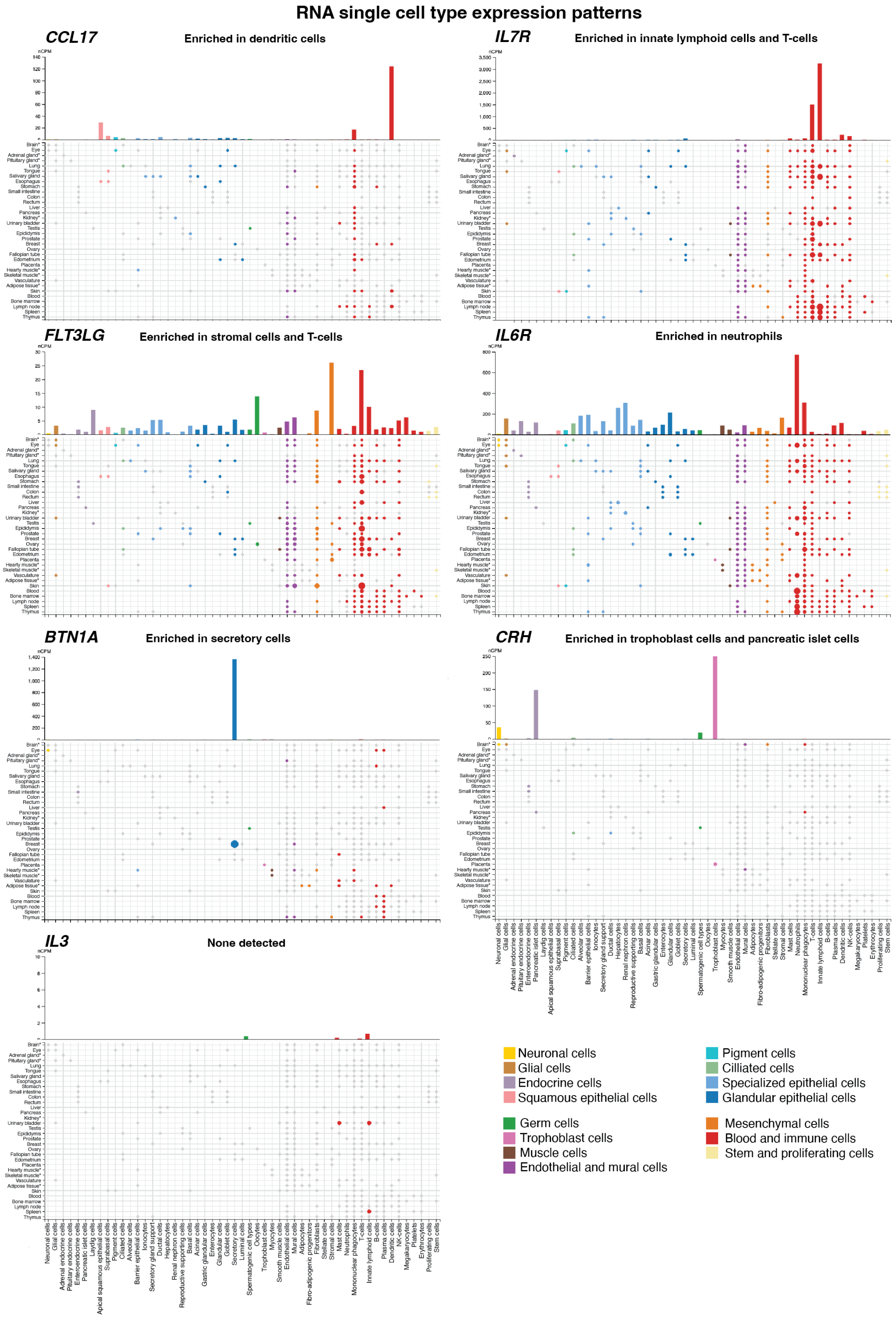


**Supplemental Figure S10. Gene expression levels of genes coding for selected extracellular proteins.** Publicly available single cell RNA-sequencing images from the Human Protein Atlas (HPA)(6-12) were utilized and merged into a single figure to check for expression levels of extracellular markers in various cell types.

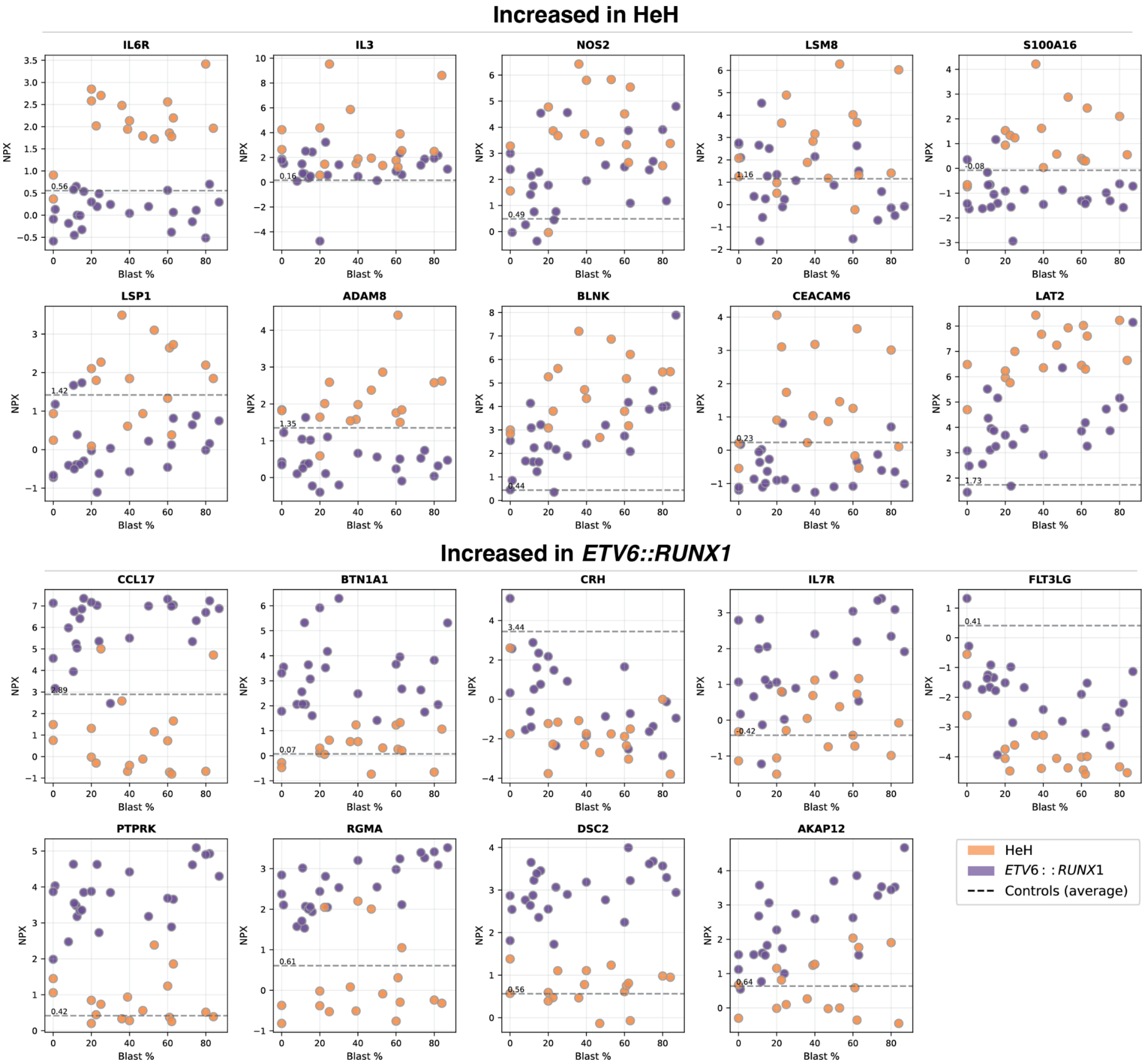


**Supplemental Figure S11. Protein abundance levels compared with peripheral blood blasts (PBB) in HeH and *ETV6::RUNX1* BCP-ALL patients.** Scatter plots showing NPX values (y-axis) of proteins increased in HeH (top) and *ETV6::RUNX1* (bottom) patients compared with PBB.

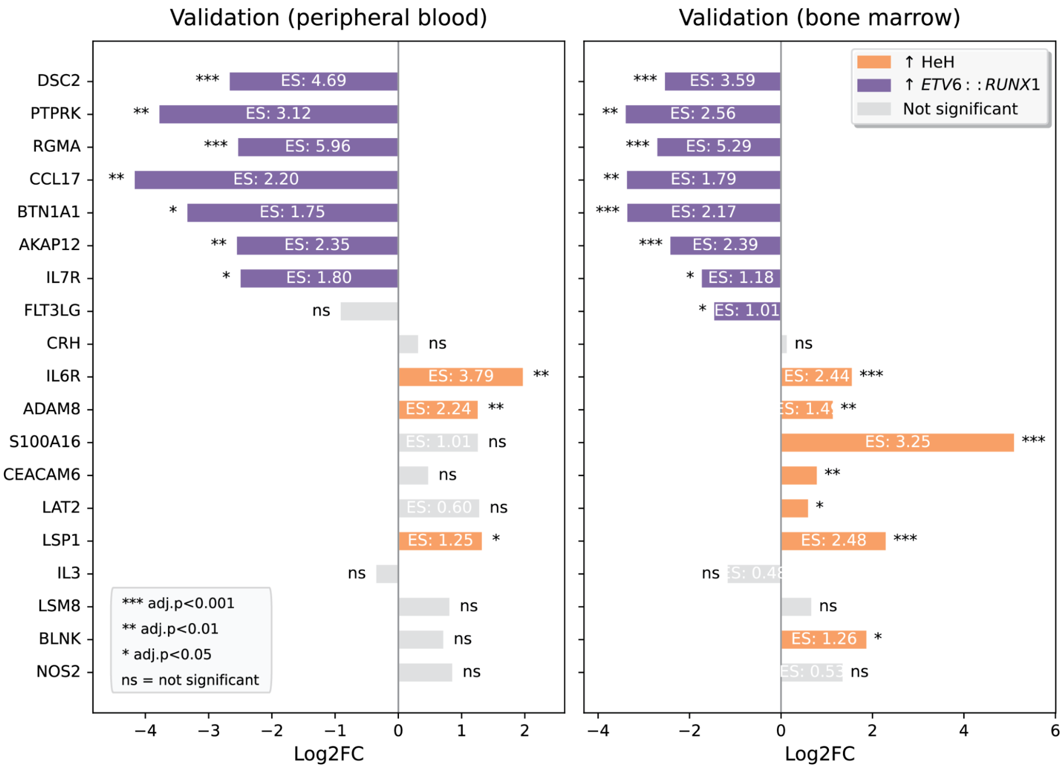


**Supplemental Figure S12. Effect size of differentially expressed proteins between HeH and *ETV6::RUNX1* in validation cohort.** Forest plots show Log2 fold change values with effect sizes (ES) for each differentially expressed protein between HeH and *ETV6::RUNX1* samples in an independent external Oink dataset of peripheral blood samples (left), and bone marrow samples (right). Data are shown for peripheral blood (n=17) and bone marrow (n=26) validation samples. Where 17 patients provided both blood and bone marrow plasma while 9 donors only provided bone marrow plasma. Yellow bars represent proteins with higher levels in HeH samples, while purple bars indicate proteins with higher levels in *ETV6::RUNX1* samples. The ES significance is annotated for each protein; ***adj.p<0.001, **adj.p<0.01, *adj.p<0.05, and ns=not significant.

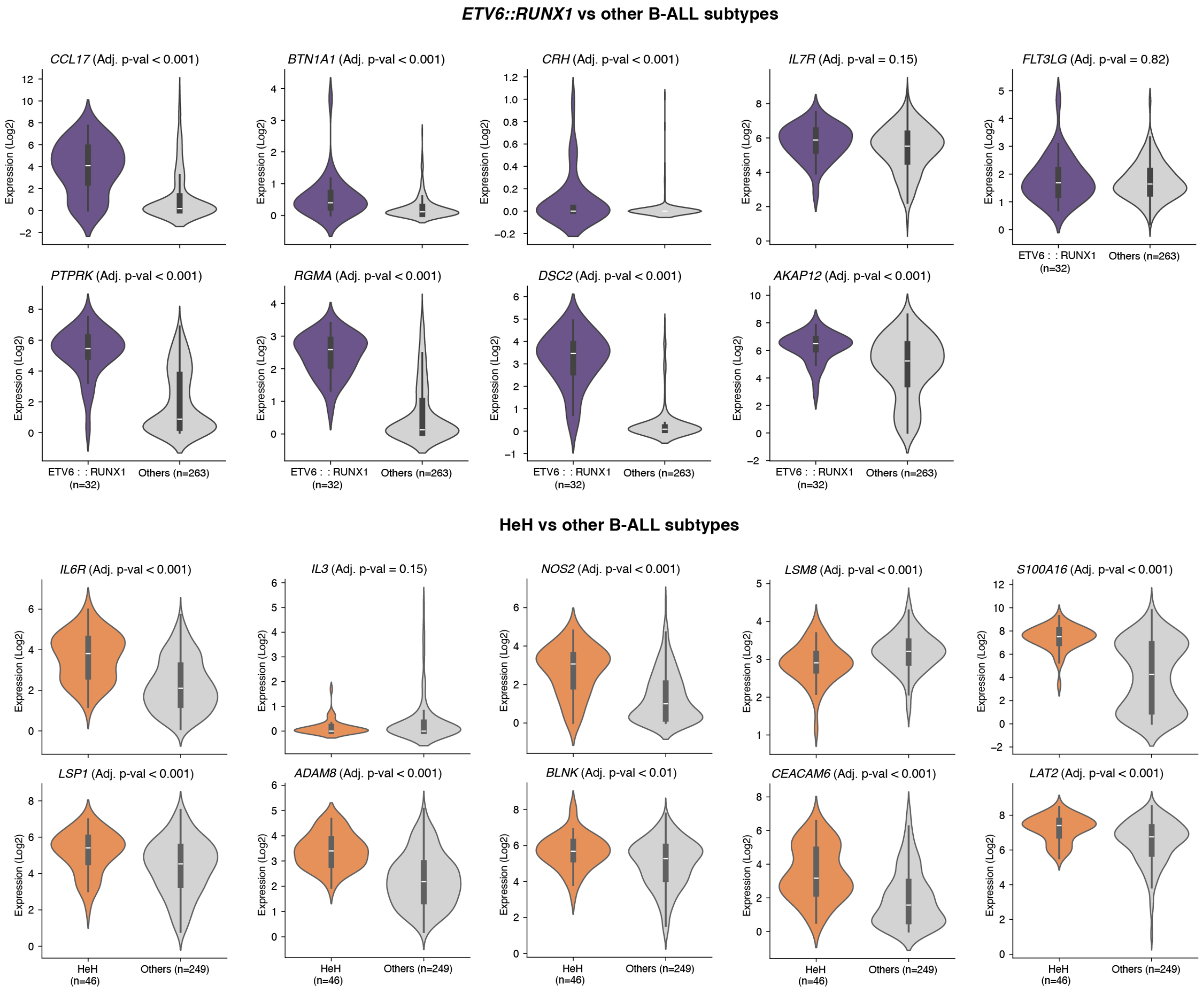


**Supplemental Figure S13. Violin plots showing genes coding for subtype-defining markers in *ETV6::RUNX1* (top) and HeH (bottom) compared with other BCP-ALL subtypes.** Each subtype (*ETV6::RUNX1* or HeH) was compared to all other BCP-ALL subtypes using a two-sided Mann–Whitney U test, and controlled FDR across the biomarkers within each subtype panel using Benjamini–Hochberg correction. RNA-seq data retrieved from Krali *et al.*, 2023 (accession number GSE227832).
